# Cohort Profile: Longitudinal population-based study of COVID-19 in UK adults (COVIDENCE UK)

**DOI:** 10.1101/2022.06.20.22276205

**Authors:** Hayley Holt, Clare Relton, Mohammad Talaei, Jane Symons, Molly R Davies, David A Jolliffe, Giulia Vivaldi, Florence Tydeman, Anne E Williamson, Paul E Pfeffer, Christopher Orton, David V Ford, Gwyneth A Davies, Ronan A Lyons, Christopher J Griffiths, Frank Kee, Aziz Sheikh, Gerome Breen, Seif O Shaheen, Adrian R Martineau

## Abstract

**Background:** Coronavirus disease 2019 (COVID-19), caused by the novel severe acute respiratory syndrome coronavirus 2 (SARS-CoV-2), is estimated to have caused more than 18 million deaths worldwide as of end-May 2022.

**Methods:** COVIDENCE UK is a longitudinal population-based study that investigates risk factors for, and impacts of, COVID-19 in UK residents aged ≥16 years. A unique feature is the capacity to support trial-within-cohort studies to evaluate interventions for prevention of COVID-19 and other acute respiratory illnesses. Participants complete a detailed online baseline questionnaire capturing self-reported information relating to their socio-demographic characteristics, occupation, lifestyle, quality of life, weight, height, longstanding medical conditions, medication use, vaccination status, diet and supplemental micronutrient intake. Follow-up on-line questionnaires capturing incident symptoms of COVID-19 and other acute respiratory infections, incident swab test-confirmed COVID-19, doses of SARS-CoV-2 vaccine received, and quality of life are completed at monthly intervals.

**Results:** The study was launched on 1^st^ May 2020 and closed to recruitment on 6^th^ October 2021. A total of 19,981 participants enrolled and consented to 5-year follow-up with medical record linkage. Their mean age was 59.1 years (range 16.0 to 94.4 years), 70.2% were female, and 93.7% identified their ethnic origin as White. Analyses conducted to date have provided key insights into risk factors for SARS-CoV-2 infection and COVID-19 disease, determinants of SARS-CoV-2 vaccine immunogenicity and efficacy, and impacts of COVID-19 on health economic outcomes. The cohort has also supported conduct of a Phase 3 randomised trial-within-cohort study (CORONAVIT) evaluating implementation of a test-and-treat approach to correcting sub-optimal vitamin D status on incidence and severity of acute respiratory infections, including COVID-19.

**Conclusions:** The COVIDENCE UK dataset represents a valuable resource containing granular information on factors influencing susceptibility to, and impacts of, COVID-19 in UK adults. Researchers wishing to access anonymised participant-level data should contacting the corresponding author for further information.

## INTRODUCTION

Coronavirus disease 2019 (COVID-19), caused by the novel severe acute respiratory syndrome coronavirus 2 (SARS-CoV-2), was first reported in Wuhan, Hubei Province, China in December 2019.^1^ The World Health Organization (WHO) declared COVID-19 a pandemic on 11^th^ March 2020.^2^ The disease is estimated to have caused more than 18 million deaths worldwide as of end-May 2022.^3^

Hospital-based studies reported risk factors for severe and fatal COVID-19 at an early stage of the pandemic.^4,5^ However, at this time we identified a need for population-based longitudinal studies to complement research conducted in secondary and tertiary care by identifying risk factors for developing predominantly mild/moderate COVID-19 that did not present to hospital. We considered this to be an important goal, both from a public health perspective (as mild/moderate disease may result in transmission to individuals who are at risk of more severe disease)^6^ and from a biological perspective (since understanding susceptibility factors can provide insights into pathogenesis).^7^ At the time, there was also a lack of longitudinal studies designed to characterise the natural history and sequelae of COVID-19 that did nor precipitate hospitalisation, and to evaluate the impact of non-hospitalised COVID-19 on adults’ physical and mental health over the longer term. Furthermore, we wished to establish a platform from which to conduct Phase 3 randomised controlled trials of non-pharmaceutical interventions for prevention of COVID-19 and other acute respiratory infections using Trials within Cohorts methodology.^8^ We therefore established a national prospective population-based longitudinal study of COVID-19 in UK adults, which we named COVIDENCE UK (https://www.qmul.ac.uk/covidence/about-the-covidence-uk-study/).

COVIDENCE UK is sponsored by, and located at, Queen Mary University of London. It was funded by Barts Charity (ref MGU0466); approved by Leicester South Research Ethics Committee (ref 20/EM/0117); and prospectively registered with ClinicalTrials.gov (NCT04330599). Enrolment opened on 1^st^ May 2020 and closed on 6^th^ October 2021.

## METHODS

Participants were invited to complete online follow-up questionnaires at monthly intervals. Cumulative totals of those who have completed monthly questionnaires, and proportions of those completing monthly questionnaires after being invited to do so are presented in Figures 1B and 1C, respectively. A total of 1,593 (8.0%) participants have withdrawn from the study to date. Table 2 compares characteristics of participants remaining in the cohort to those of participants who have withdrawn. Increased risk of withdrawal was associated with younger *vs*. older age, male *vs*. female sex, Asian/Asian British *vs*. White ethnic origin and lower *vs*. higher educational attainment.

**Figure 1.**
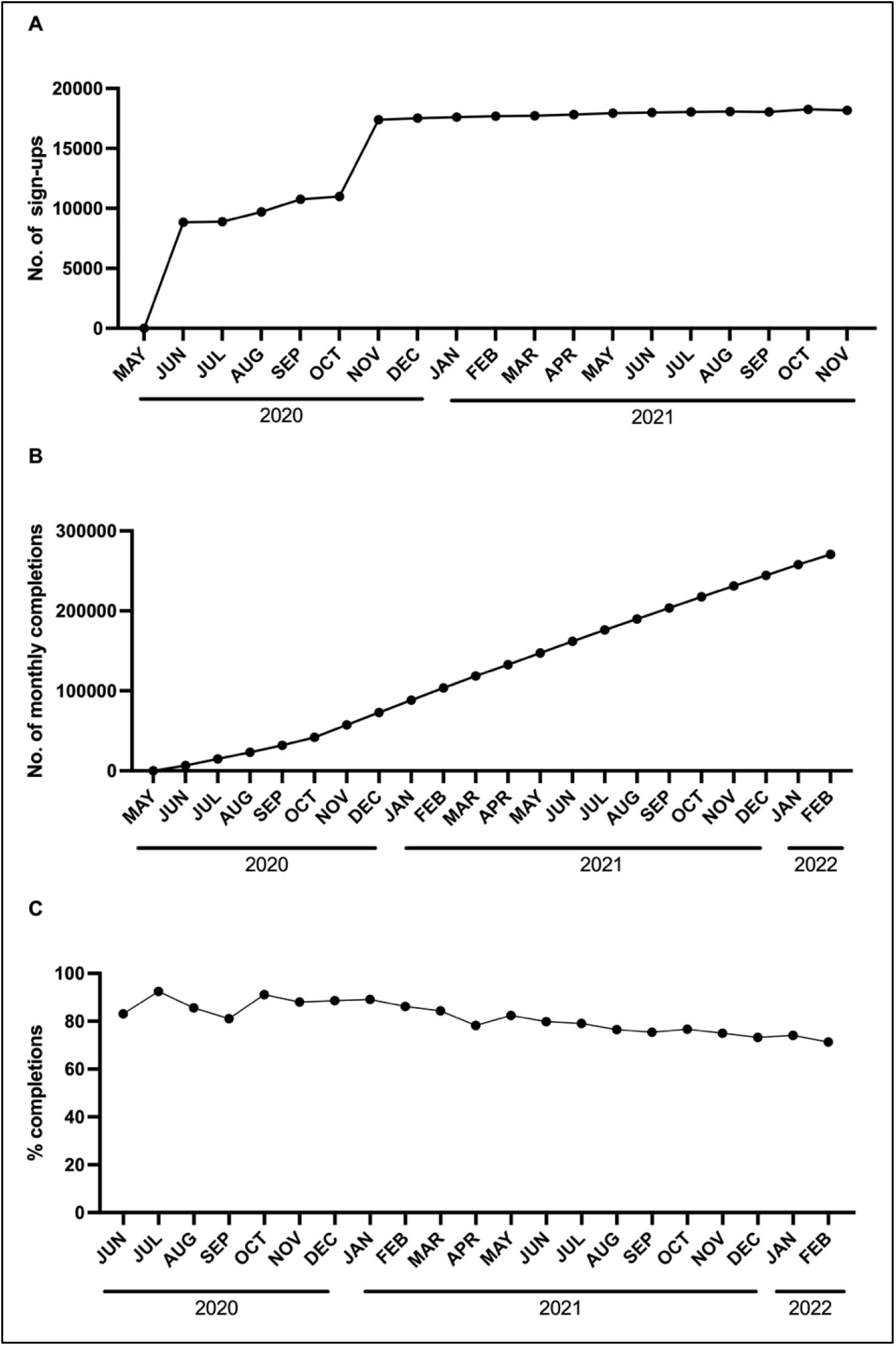
Enrolment and questionnaire completion. **A**, Cumulative number of baseline questionnaire completions, by month **B**, Cumulative number of monthly questionnaire completions, by month **C**, Proportion of participants who completed a monthly questionnaire following invitation to do so, by month

**Table 1.** Participants’ characteristics at baseline (n=19,981)

|  |  |
| --- | --- |
| **Age in years, n (%)** |  |
| 16 to <30 | 926 (4.6) |
| 30 to <40 | 1,508 (7.6) |
| 40 to <50 | 2,613 (13.1) |
| 50 to <60 | 4,581 (22.9) |
| 60 to <70 | 6,290 (31.5) |
| 70 to <80 | 3,684 (18.4) |
| ≥ 80 | 379 (1.9) |
| **Sex, n (%)** |  |
| Female | 14,028 (70.2) |
| Male | 5,953 (29.8) |
| **Ethnicity, n (%)** |  |
| White | 18,726 (93.7) |
| Mixed/Multiple/Other ethnic groups | 601 (3.0) |
| South Asian | 430 (2.2) |
| Black/African/Caribbean/Black British | 148 (0.7) |
| Data missing | 76 (0.4) |
| **Country of residence, n (%)** |  |
| England | 17,532 (87.7) |
| Northern Ireland | 389 (1.9) |
| Scotland | 1,232 (6.2) |
| Wales | 735 (3.7) |
| Data missing | 93 (0.5) |
| **Household income sufficient for basic needs, n (%)** |  |
| Yes | 18,194 (91.1) |
| Mostly / Sometimes / No | 1,656 (8.3) |
| Data missing | 131 (0.7) |
| **Housing, n (%)** |  |
| Owns own home | 11,338 (56.7) |
| Mortgage | 5,236 (26.2) |
| Privately Renting | 1,588 (7.9) |
| Renting from council | 781 (3.9) |
| Others | 932 (4.7) |
| Data missing | 106 (0.5) |
| **Highest educational level attained, n (%)** |  |
| Primary/Secondary | 2,192 (11.0) |
| Higher/further (A levels) | 2,955 (14.8) |
| College | 8,769 (43.8) |
| Post-graduate | 5,947 (29.8) |
| Data missing | 118 (0.6) |
| **Occupational status, n (%)** |  |
| Employed | 7,458 (37.3) |
| Self-employed | 1,856 (9.3) |
| Retired | 8,316 (41.6) |
| Furloughed | 451 (2.3) |
| Unemployed | 394 (2.0) |
| Student | 438 (2.2) |
| Other | 988 (5.0) |
| Data missing | 80 (0.4) |
| **Frontline worker, n (%)** |  |
| No | 15,802 (79.1) |
| Other frontline worker | 2,255 (11.3) |
| Health or social care worker | 1,812 (9.1) |
| Data missing | 112 (0.6) |
| **Body mass index, n (%)** |  |
| <25, kg/m2 | 9,501 (47.6) |
| 25-30, kg/m2 | 6,343 (31.7) |
| >30, kg/m2 | 4,027 (20.2) |
| Data missing | 110 (0.6) |
| **Self-reported general health, n (%)** |  |
| Excellent | 3,683 (18.4) |
| Very good | 7,444 (37.3) |
| Good | 5,308 (26.6) |
| Fair | 2,369 (11.9) |
| Poor | 883 (4.4) |
| Data missing | 294 (1.5) |
| **Tobacco smoking history, n (%)** |  |
| Never-smoker | 10,978 (54.9) |
| Ex-smoker | 7,645 (38.3) |
| Current smoker | 1,220 (6.1) |
| Data missing | 138 (0.7) |
| **E-cigarette use, n (%)** |  |
| Never-user | 18,302 (91.6) |
| Ex-user | 780 (3.9) |
| Current user | 700 (3.5) |
| Data missing | 199 (1.0) |
| **Alcohol consumption in week prior to questionnaire completion, n (%)** |  |
| None | 6,002 (30.0) |
| 1-7 units | 6,849 (34.3) |
| 8-14 units | 3,737 (18.7) |
| 15-21 units | 1,779 (8.9) |
| 22-28 units | 817 (4.1) |
| >28 units | 639 (3.2) |
| Data missing | 158 (0.8) |

**Table 2.** Characteristics of participants who have *vs*. have not withdrawn from follow-up.

| Characteristic | Categories | N (%) who have not withdrawn (n=18,388)<sup>1</sup> | N (%) who have withdrawn (n=1,593)<sup>1</sup> |
| --- | --- | --- | --- |
| Age, years | 16 to <30 | 794 (4.3) | 132 (8.3) |
|  | 30 to <40 | 1339 (7.3) | 169 (10.6) |
|  | 40 to <50 | 2379 (12.9) | 234 (14.7) |
|  | 50 to <60 | 4263 (23.2) | 318 (20.0) |
|  | 60 to <70 | 5892 (32.0) | 398 (25.0) |
|  | 70 to <80 | 3391 (18.4) | 293 (18.4) |
|  | ≥ 80 | 330 (1.8) | 49 (3.1) |
| Sex | Female | 13073 (71.1) | 955 (59.9) |
|  | Male | 5315 (28.9) | 638 (40.1) |
| Ethnicity | White | 17266 (93.9) | 1460 (91.7) |
|  | Mixed/Multiple/Other ethnic groups | 542 (2.9) | 59 (3.7) |
|  | Asian / Asian British | 374 (2.0) | 56 (3.5) |
|  | Black/African/Caribbean/Black British | 130 (0.7) | 18 (1.1) |
|  | Data missing | 76 (0.4) | 0 (0.00) |
| Highest educational level attained | Primary/Secondary | 1932 (10.5) | 260 (16.3) |
|  | Higher/further (A levels) | 2676 (14.6) | 279 (17.5) |
|  | College or University | 8106 (44.1) | 663 (41.6) |
|  | Post-graduate | 5157 (28.0) | 390 (24.5) |
|  | Data missing | 117 (0.6) | 1 (0.06) |
| Household income sufficient to cover basic needs | Yes | 16752 (91.1) | 1442 (90.5) |
|  | Mostly / Sometimes / No | 1505 (8.2) | 151 (9.5) |
|  | Data missing | 131 (0.7) | 0 (0.00) |
| Geographical location | England | 16153 (87.8) | 1379 (86.6) |
|  | Northern Ireland | 372 (2.0) | 39 (2.4) |
|  | Scotland | 1186 (6.4) | 101 (6.3) |
|  | Wales | 699 (3.8) | 70 (4.4) |
|  | Data missing | 89 (0.5) | 4 (0.3) |

1, as of 21<sup>st</sup> April 2022.

Table 3 presents a high-level summary of key variables for which data are available, along with details relating to the sources of these data. Full details of questions in baseline and monthly questionnaires are presented in the Supplementary Appendix.

**Table 3.**
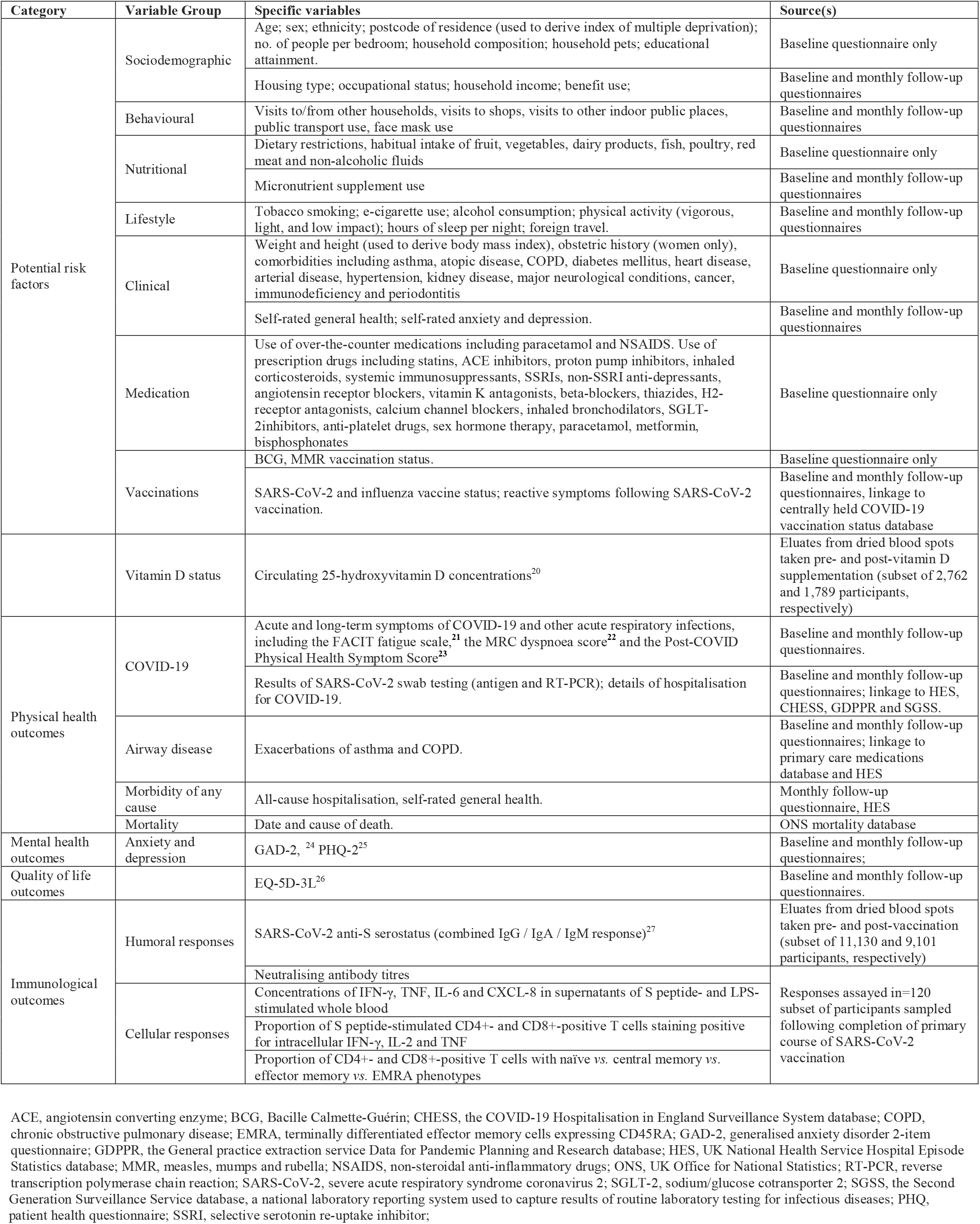
Variables captured and relevant data sources.

## RESULTS

Baseline characteristics of the 19,981 participants who enrolled in the cohort and completed the baseline questionnaire are presented in Table 1: mean age was 59.1 years (range 16.0 to 94.4 years), 70.2% were female, 87.7% lived in England, and 93.7% identified their ethnic origin as White. Participants were recruited following a national media campaign with publicity relating to the study appearing in print and online newspapers, radio, television, social media and online advertising; the timeline for participant accrual is illustrated in Figure 1A. Heatmaps illustrate a high degree of overlap between participants’ area of residence (Figure 2A) and the location of COVID-19 cases notified to UK public health authorities (Figure 2B).

**Figure 2.**
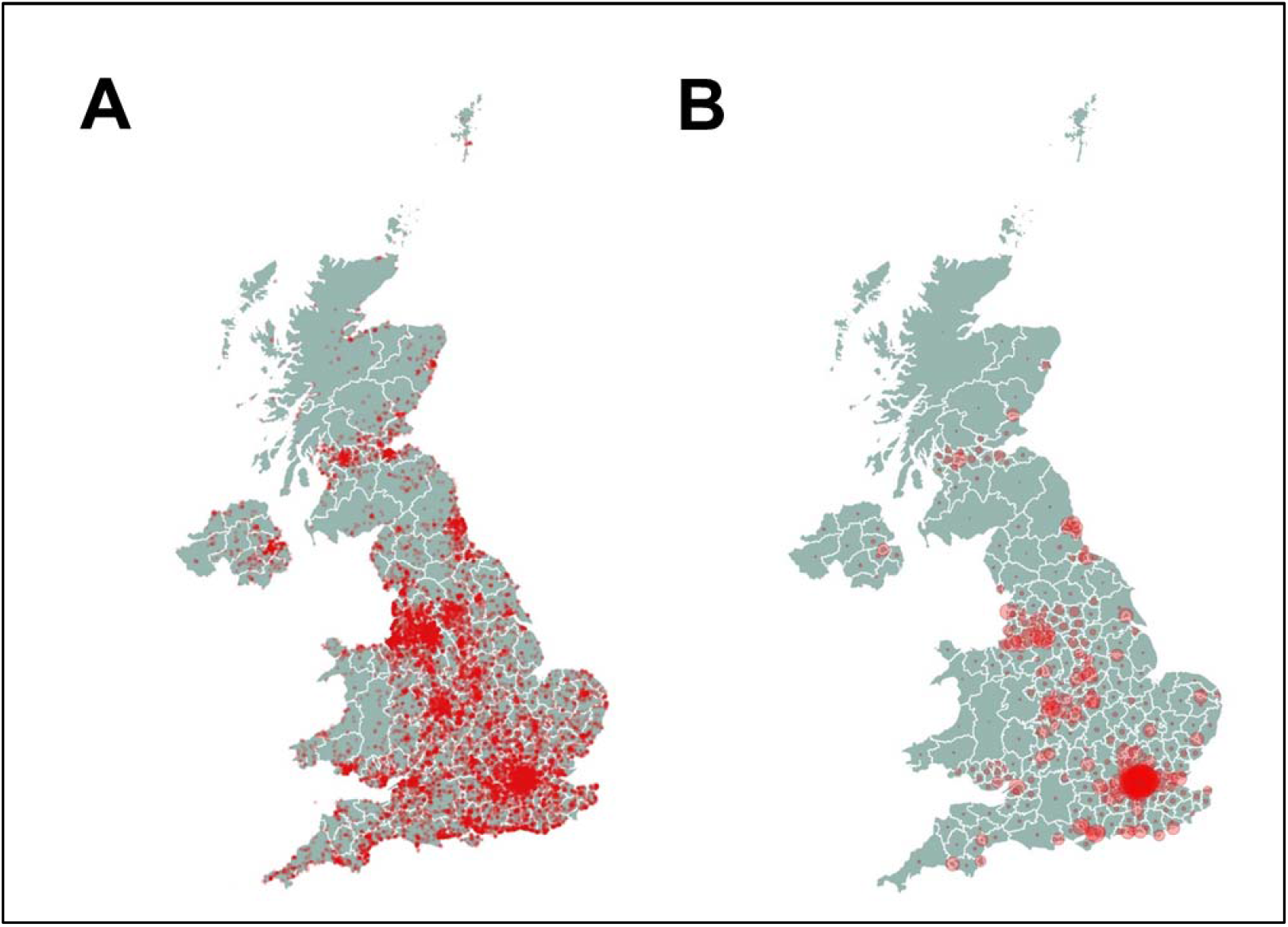
Geographical location of participants and COVID-19 cases. **A**, heatmap of COVIDENCE UK participants’ location of residence by postcode. **B**, heatmap of cumulative COVID-19 notifications in the UK, by postcode, from https://coronavirus.data.gov.uk/details/download (download dated June 10, 2022).

Key findings to date include the following:

- Asian/Asian British ethnicity was associated with increased odds both of serologically confirmed SARS-CoV-2 infection and of developing swab test-confirmed COVID-19, after rigorous adjustment for potential confounders including socioeconomic status, presence of comorbidities and multiple social and environmental factors influencing exposure to SARS-CoV-2.^7,9^ Participants of Asian/Asian British origin also had higher convalescent titres of combined IgG, IgA and IgM antibodies to the SARS-CoV-2 spike protein following infection (after adjustment for multiple potential confounders including disease severity) and higher titres of the same antibodies after SARS-CoV-2 vaccination (after adjustment for multiple potential confounders including pre-vaccination anti-S titres).^9,10^ Taken together, these findings highlight the need to investigate potential biological determinants of ethnic variation in susceptibility to, and severity of, COVID-19.
- Similarly, higher body mass index was found to associate independently with increased susceptibility to SARS-CoV-2 infection and COVID-19, and higher titres of anti-S antibodies following COVID-19 and vaccination against SARS-CoV-2.^7,9,10^ The mechanisms underlying these associations are also worthy of investigation.
- By contrast, we found that several factors associating with COVID-19 severity including age, sex and presence of comorbidities including ischaemic heart disease, hypertension and diabetes mellitus did not associate with susceptibility to developing COVID-19.^7^ The degree of overlap between factors influencing susceptibility to COVID-19 *vs*. disease severity therefore appears to be limited (Figure 3).
- Analyses of post-vaccination serology data showed that higher post-vaccination titres of anti-S antibodies associated with self-report of reactive symptoms following SARS-CoV-2 vaccination^11^ and were a correlate of protection against breakthrough COVID-19.^12^ We also identified several factors associating with lower anti-S antibody titres following a primary course of vaccination against SARS-CoV-2, including vaccination with ChAdOx1 nCoV-19 (Oxford AstraZeneca, ChAdOx1) *vs*. BNT162b2 (Pfizer), a shorter interval between vaccine doses, poor *vs*. excellent self-rated general health, immunodeficiency and use of immunosuppressant medications. Among a subset of 247 participants who were seronegative following their primary course of vaccination, administration of a single booster dose of mRNA vaccine was effective in achieving seroconversion in all but eight individuals, all of whom had a known immunodeficiency or were taking immunosuppressant medication.^10^
- Analyses investigating incidence of SARS-CoV-2 infection in vaccinated participants showed that post-vaccination titres of anti-S antibodies were a correlate of protection against breakthrough COVID-19,^12^ and that risk of breakthrough disease was independently associated with younger *vs*. older age, lower *vs*. higher educational attainment, administration of ChAdOx1 *vs*. BNT182b2, and more *vs*. less frequent visits to indoor public places.^13^
- A phase 3 trial-within-cohort study conducted in a subset of 6,200 COVIDENCE UK participants from December 2020 to June 2021 showed that implementation of a test-and-treat approach to correction of sub-optimal vitamin D status did not result in reduced incidence or severity of COVID-19.^14^
- Health economic analyses of data from the cohort have demonstrated an independent association between incident COVID-19 and subsequently increased risk of reporting long-term sickness absence from work and household income being insufficient to meet basic needs.^15^ Given that socio-economic disadvantage is recognised to increase the risk of developing COVID-19,^16^ this finding raises the prospect that COVID-19 may generate a vicious cycle of impaired health and poor economic outcomes.

**Figure 3.**
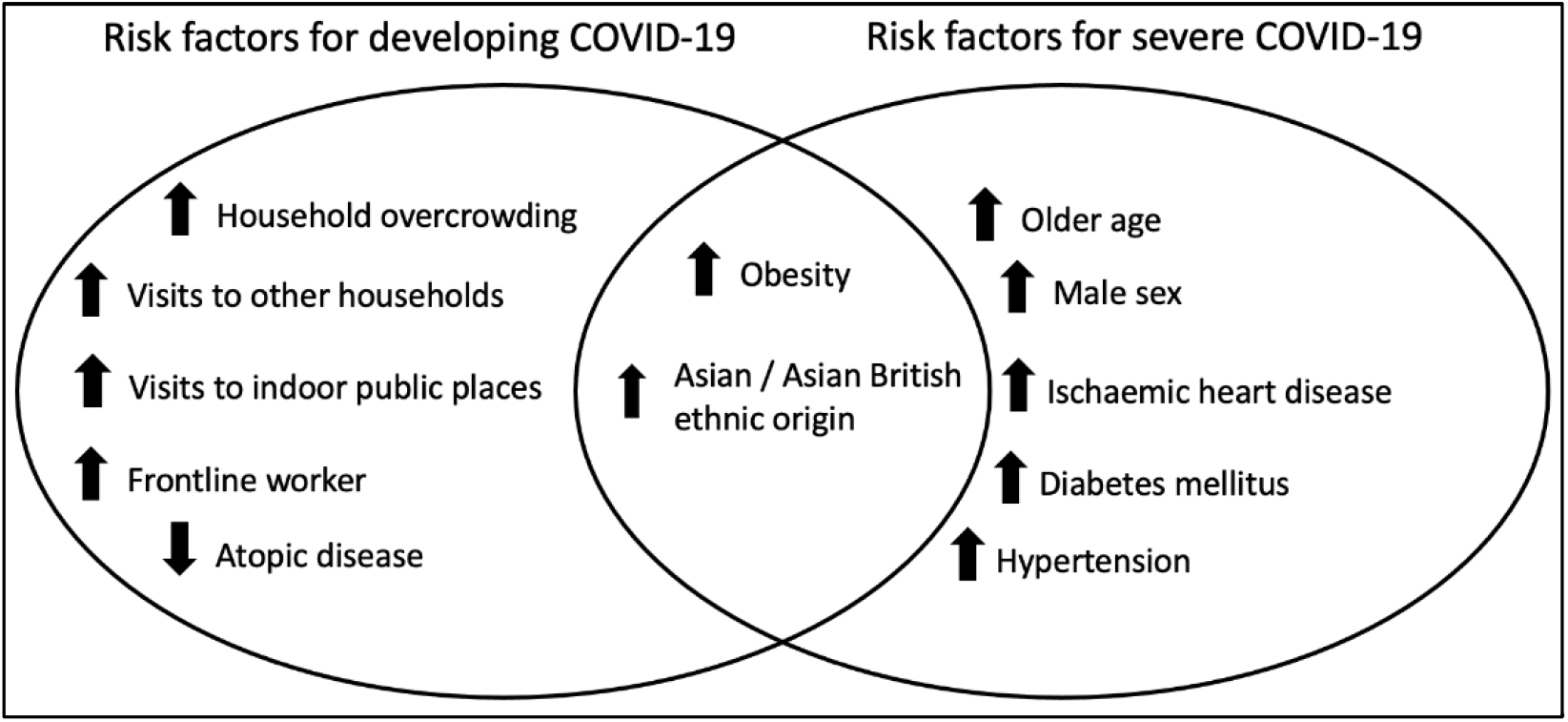
Risk factors for developing COVID-19 identified by COVIDENCE UK compared to risk factors for severe or fatal disease. Upward/downward arrows indicate increased/decreased risk, respectively.

## DISCUSSION

### What are the main strengths and weaknesses?

A major strength is that COVIDENCE UK was established specifically in response to the SARS-CoV-2 outbreak; thus, in contrast to previously established cohort studies, our questionnaires were tailored to capture potential risk factors and incident symptoms of COVID-19. Another strength of our study is that participants have given consent to be followed up for five years with full medical record linkage; thus if they die or fail to complete follow-up questionnaires, health outcomes will continue to be captured. Monthly follow-up reduces potential issues associated with poor recall of symptoms and health events that might arise with longer intervals between questionnaires. Our efforts to maintain participant engagement via monthly webinars, coupled with high degrees of public concern about the pandemic, are reflected in our low withdrawal rate (8%). In contrast to studies focusing exclusively on disease presenting to hospital, COVIDENCE UK is particularly well-positioned to identify risk factors for developing mild and moderate COVID-19, providing important insights into factors affecting disease transmission. Finally, the capacity of the cohort to support trial-within-cohort studies^14^ allows us to conduct phase 3 randomised controlled trials of interventions to prevent COVID-19 and other acute respiratory illnesses rapidly and efficiently.

The principal weakness of the cohort relates to under-representation of certain population groups, including younger adults, men, people with lower educational attainment and ethnic minorities: this may introduce collider bias,^17^ reduce the generalisability of our findings, and limit our power to detect risk factors for COVID-19 in these groups. However, we highlight that representativeness is not necessarily a barrier to addressing aetiological questions.^18,19^ Moreover, other groups at heightened risk of COVID-19 such as older adults and people with comorbidities are over-represented in the cohort, making us well placed to study them.

### Can I get hold of the data? Where can I find out more?

Data can be accessed by contacting the corresponding author. Study findings will be presented at international conferences and published in peer-reviewed journals.

## Data Availability

All data produced in the present work are contained in the manuscript

## Author Contributions

HH and ARM wrote the first draft of the paper. All co-authors reviewed and contributed to the writing of the manuscript.

## Acknowledgements

We thank all COVIDENCE UK participants, and all our funders for their generous support.

## References

1. Wang C, Horby PW, Hayden FG, Gao GF. A novel coronavirus outbreak of global health concern. Lancet 2020; 395(10223): 470–3. 10.1016/S0140-6736(20)30185-9.

2. Cucinotta D, Vanelli M. WHO Declares COVID-19 a Pandemic. Acta Biomed 2020; 91(1): 157–60. 10.23750/abm.v91i1.9397.

3. Covid-19 Excess Mortality Collaborators. Estimating excess mortality due to the COVID-19 pandemic: a systematic analysis of COVID-19-related mortality, 2020-21. Lancet 2022; 399(10334): 1513–36. 10.1016/S0140-6736(21)02796-3.

4. Zhou F, Yu T, Du R, Fan G, Liu Y, Liu Z, Xiang J, Wang Y, Song B, Gu X, Guan L, Wei Y, Li H, Wu X, Xu J, Tu S, Zhang Y, Chen H, Cao B. Clinical course and risk factors for mortality of adult inpatients with COVID-19 in Wuhan, China: a retrospective cohort study. Lancet 2020; 395(10229): 1054–62. 10.1016/S0140-6736(20)30566-3.

5. Clift AK, Coupland CAC, Keogh RH, Diaz-Ordaz K, Williamson E, Harrison EM, Hayward A, Hemingway H, Horby P, Mehta N, Benger J, Khunti K, Spiegelhalter D, Sheikh A, Valabhji J, Lyons RA, Robson J, Semple MG, Kee F, Johnson P, Jebb S, Williams T, Hippisley-Cox J. Living risk prediction algorithm (QCOVID) for risk of hospital admission and mortality from coronavirus 19 in adults: national derivation and validation cohort study. BMJ 2020; 371: m3731. 10.1136/bmj.m3731.

6. Boehmer TK, DeVies J, Caruso E, van Santen KL, Tang S, Black CL, Hartnett KP, Kite-Powell A, Dietz S, Lozier M, Gundlapalli AV. Changing Age Distribution of the COVID-19 Pandemic - United States, May-August 2020. MMWR Morb Mortal Wkly Rep 2020; 69(39): 1404–9. 10.15585/mmwr.mm6939e1.

7. Holt H, Talaei M, Greenig M, Zenner D, Symons J, Relton C, Young KS, Davies MR, Thompson KN, Ashman J, Rajpoot SS, Kayyale AA, El Rifai S, Lloyd PJ, Jolliffe D, Timmis O, Finer S, Iliodromiti S, Miners A, Hopkinson NS, Alam B, Lloyd-Jones G, Dietrich T, Chapple I, Pfeffer PE, McCoy D, Davies G, Lyons RA, Griffiths C, Kee F, Sheikh A, Breen G, Shaheen SO, Martineau AR. Risk factors for developing COVID-19: a population-based longitudinal study (COVIDENCE UK). Thorax 2021. 10.1136/thoraxjnl-2021-217487.

8. Relton C, Torgerson D, O’Cathain A, Nicholl J. Rethinking pragmatic randomised controlled trials: introducing the “cohort multiple randomised controlled trial” design. BMJ 2010; 340: c1066. 10.1136/bmj.c1066.

9. Talaei M, Faustini S, Holt H, Jolliffe DA, Vivaldi G, Greenig M, Perdek N, Maltby S, Bigogno CM, Symons J, Davies GA, Lyons RA, Griffiths CJ, Kee F, Sheikh A, Richter AG, Shaheen SO, Martineau AR. Determinants of pre-vaccination antibody responses to SARS-CoV-2: a population-based longitudinal study (COVIDENCE UK). BMC Medicine 2022; 20(1): 87. 10.1186/s12916-022-02286-4.

10. Jolliffe DA, Faustini SE, Holt H, Perdek N, Maltby S, Talaei M, Greenig M, Sheikh A, Lyons RA, Kee F, Martineau AR. Determinants of Antibody Responses to Two Doses of ChAdOx1 nCoV-19 or Bnt162b2 and a Subsequent Booster Dose of BNT162b2 or mRNA-1273: Population-Based Cohort Study (COVIDENCE UK). Preprints with The Lancet, 2022.http://dx.doi.org/10.2139/ssrn.4031570.

11. Holt H, Jolliffe DA, Talaei M, Faustini SE, Vivaldi G, Davies GA, Lyons RA, Griffiths CJ, Kee F, Sheikh A, Richter AG, Shaheen SO, Martineau AR. Incidence, determinants and serological correlates of reactive symptoms following SARS-CoV-2 vaccination: a population-based longitudinal study (COVIDENCE UK). Research Square 2022.https://doi.org/10.21203/rs.3.rs-1692845/v1.

12. Vivaldi G, Jolliffe DA, Faustini SE, Holt H, Perdek N, Talaei M, Tydeman F, Chambers ES, Cai W, Li W, Gibbons J, Pade C, McKnight A, Shaheen SO, Richter AG, Martineau AR. Correlation between post-vaccination titres of IgG, IgA and IgM anti-Spike antibodies and protection against breakthrough SARS-CoV-2 infection: a population-based longitudinal study (COVIDENCE UK). MedRxiv 2022.https://doi.org/10.1101/2022.02.11.22270667

13. Vivaldi G, Jolliffe DA, Holt H, Tydeman F, Talaei M, Davies GA, Lyons RA, Griffiths CJ, Kee F, Sheikh A, Shaheen SO, Martineau AR. Risk factors for SARS-CoV-2 infection after primary vaccination with ChAdOx1 nCoV-19 or BNT1262b2 and after booster vaccination with BNT1262b2 or mRNA-1273: a population-based cohort study (COVIDENCE UK). MedRxiv 2022.https://doi.org/10.1101/2022.03.11.22272276.

14. Jolliffe DA, Holt H, Greenig M, Talaei M, Perdek N, Pfeffer P, Maltby S, Symons J, Barlow NL, Normandale A, Garcha R, Richter AG, Faustini SE, Orton C, Ford DV, Lyons RA, Davies GA, Kee F, Griffiths CJ, Norrie J, Sheikh A, Shaheen SO, Relton C, Martineau AR. Vitamin D Supplements for Prevention of Covid-19 or other Acute Respiratory Infections: a Phase 3 Randomized Controlled Trial (CORONAVIT). MedRxiv 2022.https://doi.org/10.1101/2022.03.22.22271707.

15. Williamson AE, Tydeman F, Miners A, Pyper K, Martineau AR. Acute and long-term impacts of COVID-19 on economic vulnerability: a population-based longitudinal study (COVIDENCE UK). MedRxiv 2022.https://doi.org/10.1101/2022.03.03.22271835.

16. Patel AP, Paranjpe MD, Kathiresan NP, Rivas MA, Khera AV. Race, socioeconomic deprivation, and hospitalization for COVID-19 in English participants of a national biobank. Int J Equity Health 2020; 19(1): 114. 10.1186/s12939-020-01227-y.

17. Griffith GJ, Morris TT, Tudball MJ, Herbert A, Mancano G, Pike L, Sharp GC, Sterne J, Palmer TM, Davey Smith G, Tilling K, Zuccolo L, Davies NM, Hemani G. Collider bias undermines our understanding of COVID-19 disease risk and severity. Nature Communications 2020; 11(1): 5749. 10.1038/s41467-020-19478-2.

18. Doll R, Hill AB. Mortality in Relation to Smoking: Ten Years’ Observations of British Doctors. Br Med J 1964; 1(5396): 1460–7 CONCL. 10.1136/bmj.1.5396.1460.

19. Rothman KJ, Gallacher JE, Hatch EE. Why representativeness should be avoided. Int J Epidemiol 2013; 42(4): 1012–4. 10.1093/ije/dys223.

20. Shea RL, Berg JD. Self-administration of vitamin D supplements in the general public may be associated with high 25-hydroxyvitamin D concentrations. Ann Clin Biochem 2017; 54(3): 355–61. 10.1177/0004563216662073.

21. Yellen SB, Cella DF, Webster K, Blendowski C, Kaplan E. Measuring fatigue and other anemia-related symptoms with the Functional Assessment of Cancer Therapy (FACT) measurement system. J Pain Symptom Manage 1997; 13(2): 63–74. 10.1016/s0885-3924(96)00274-6.

22. Fletcher CM. Standardised questionnaire on respiratory symptoms: a statement prepared and approved by the MRC Committee on the Aetiology of Chronic Bronchitis (MRC breathlessness score). BMJ 1960; 2: 1665.

23. Taylor RR, Trivedi B, Patel N, Singh R, Ricketts WM, Elliott K, Yarwood M, White V, Hylton H, Allen R, Thomas G, Kapil V, McGuckin R, Pfeffer PE. Post-COVID symptoms reported at asynchronous virtual review and stratified follow-up after COVID-19 pneumonia. Clin Med (Lond) 2021. 10.7861/clinmed.2021-0037.

24. Kroenke K, Spitzer RL, Williams JB, Monahan PO, Lowe B. Anxiety disorders in primary care: prevalence, impairment, comorbidity, and detection. Ann Intern Med 2007; 146(5): 317–25. 10.7326/0003-4819-146-5-200703060-00004.

25. Kroenke K, Spitzer RL, Williams JB. The Patient Health Questionnaire-2: validity of a two-item depression screener. Med Care 2003; 41(11): 1284–92. 10.1097/01.MLR.0000093487.78664.3C.

26. EuroQol G. EuroQol--a new facility for the measurement of health-related quality of life. Health Policy 1990; 16(3): 199–208. 10.1016/0168-8510(90)90421-9.

27. Morley GL, Taylor S, Jossi S, Perez-Toledo M, Faustini SE, Marcial-Juarez E, Shields AM, Goodall M, Allen JD, Watanabe Y, Newby ML, Crispin M, Drayson MT, Cunningham AF, Richter AG, O’Shea MK. Sensitive Detection of SARS-CoV-2-Specific Antibodies in Dried Blood Spot Samples. Emerging Infectious Diseases 2020; 26(12): 2970–3. 10.3201/eid2612.203309.

