## Supplementary Appendix COVIDENCE UK cohort profile for "Cohort Profile: Longitudinal population-based study of COVID-19 in UK adults (COVIDENCE UK)" (from another version of this preprint: version 1, 2022-06-20)

### COVIDENCE UK Baseline Questionnaire

Welcome to your one-time baseline questionnaire for the COVIDENCE UK study.

Before you start, please make sure that you have any hospital letters or prescriptions handy, as we’ll be asking for very precise details about any medical conditions you have or treatments you’ve been prescribed. The questionnaire can take up to 60 minutes to complete, although most people will be done in 30 minutes.

Please select your answers carefully. Questions that only require one answer will automatically forward you to the next question. If you feel that there is important additional information that has not been captured by our questionnaire, you can enter this into the free text box provided at the end of the survey.

First we will ask about your height and weight.

End of Block: COVIDENCE_Intro

Start of Block: COVIDENCE_Demographics

| 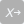 | 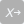 |
| --- | --- |

Q1 Please select your preferred unit of measurement for height:

- Feet/inches (0)
- Centimetres (1)

| Page Break |
| --- |

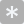

Q2 What is your **current** height?  (if you are unsure, please put your best estimate)

*If you decide you would like to go back and answer in a different unit, please make sure to remove your response from this box.*

Q1 = Feet/inches

- Feet (ft) (1) ________________________________________________

Q1 = Feet/inches

- Inches (in) (2) ________________________________________________

Q1 = Centimetres

- Centimetres (cm) (4) ________________________________________________

| Page Break |
| --- |

| 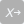 | 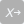 |
| --- | --- |

Q3 Please select your preferred unit of measurement for weight:

- Stones/pounds (0)
- Kilograms (1)

| Page Break |
| --- |

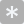

Q4 What is your **current** weight?

*If you decide you would like to go back and answer in a different unit, please make sure to remove your response from this box.*

Q3 = Stones/pounds

- Stones (sts) (1) ________________________________________________

Q3 = Stones/pounds

- Pounds (lbs) (2) ________________________________________________

Q3 = Kilograms

- Kilograms (kg) (3) ________________________________________________

| Page Break |
| --- |

| 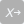 | 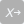 |
| --- | --- |

Q5 What is your ethnic origin?

- White (0)
- Mixed / Multiple ethnic groups (1)
- Asian / Asian British (2)
- Black / African / Caribbean / Black British (3)
- Arab (4)
- Other Ethnic Group (5)

| Page Break |
| --- |

Display This Question:

If Q5 = White

| 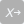 | 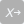 |
| --- | --- |

Q6 Please tell us more about your ethnic origin

- English / Welsh / Scottish / Northern Irish / British (0)
- Irish (1)
- Gypsy or Irish Traveller (2)
- Any other white background (3)

| Page Break |
| --- |

Display This Question:

If Q5 = Mixed / Multiple ethnic groups

| 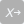 | 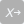 |
| --- | --- |

Q7 Please tell us more about your ethnic origin:

- White and Black Caribbean (0)
- White and Black African (1)
- White and Asian (2)
- Any other Mixed / Multiple ethnic backgrounds (3)

| Page Break |
| --- |

Display This Question:

If Q5 = Asian / Asian British

| 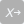 | 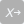 |
| --- | --- |

Q8 Please tell us more about your ethnic origin:

- Indian (0)
- Pakistani (1)
- Bangladeshi (2)
- Chinese (3)
- Any other Asian background (4)

| Page Break |
| --- |

Display This Question:

If Q5 = Black / African / Caribbean / Black British

| 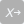 | 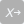 |
| --- | --- |

Q9 Please tell us more about your ethnic origin:

- African (0)
- Caribbean (1)
- Any other Black / African / Caribbean background (2)

| Page Break |
| --- |

| 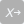 | 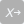 |
| --- | --- |

Q10 Were you born in the UK?

- Yes (1)
- No (0)

| Page Break |
| --- |

Q11 The next section asks about your employment status, education, and household.

| 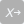 | 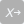 |
| --- | --- |

Q12 Which of the following best describes your current occupational status?

- Employed (0)
- Self-employed (1)
- Retired (2)
- Furloughed (3)
- Unemployed (4)
- Not working due to sickness/ disability or illness (5)
- Never employed (6)
- Student (7)
- Other	(8)

| Page Break |
| --- |

Display This Question:

If Q12 = Unemployed

| 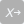 | 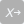 |
| --- | --- |

Q13 How long have you been unemployed?

- Less than 1 month (0)
- 1-3 months (1)
- 3-6 months (2)
- 6-12 months (3)
- 1-4 years (4)
- 5 years or more (5)

| Page Break |
| --- |

Q14 The following questions refer to your current main job, or (if you are not working now) to your last main job. Please select one box per question only.

| 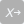 | 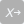 |
| --- | --- |

Q15 Do (did) you work as an employee or are (were) you self-employed?

- Employee (0)
- Self-employed with employees (1)
- Self-employed/ freelance without employees (2)
- Other (e.g. full time student, never employed) (3)

| Page Break |
| --- |

Display This Question:

If Q15 = Self-employed with employees

Q16 Indicate below how many people you employ (employed):

________________________________________________________________

| Page Break |
| --- |

Display This Question:

If Q15 = Employee

| 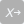 | 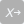 |
| --- | --- |

Q17
Do (did) you supervise any other employees?

*A supervisor or foreman is responsible for overseeing the work of other employees on a day-to-day basis.*

- Yes (1)
- No (0)

| Page Break |
| --- |

| 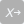 | 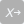 |
| --- | --- |

Q18 Which of the following best describes the sort of work you do? 
If you are not working now, please select a box to show what you did in your last job.

- Modern professional occupations *Such as: teacher – nurse – physiotherapist – social worker – welfare officer – artist – musician – police officer (sergeant or above) – software designer* (0)
- Clerical and intermediate occupations *Such as: secretary – personal assistant – clerical worker – office clerk – call centre agent – nursing auxiliary – nursery nurse* (1)
- Senior managers or administrators (usually responsible for planning, organizing, and co-ordinating work and for finance) *Such as: finance manager – chief executive* (2)
- Technical and craft occupations *Such as: motor mechanic – fitter – inspector – plumber –printer– tool maker–electrician– gardener– train driver* (3)
- Semi-Routine manual and service occupations *Such as: postal worker–machine operative– security guard–caretaker–farm-worker–catering assistant–receptionist–sales assistant* (4)
- Routine manual and service occupations *Such as: HGV driver–van driver–cleaner–porter–packer–sewing machinist– messenger–labourer–waiter/waitress–bar staff* (5)
- Middle or junior managers *Such as: office manager–retail manager–bank manager–restaurant manager–warehouse manager–publican* (6)
- Traditional professional occupations *Such as: accountant–solicitor–medical practitioner–scientist–civil/mechanical engineer* (7)
- Student (8)
- Never employed or long term (more than 5 years) unemployed (9)

| Page Break |
| --- |

| 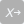 | 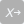 |
| --- | --- |

Q19 Are you a ‘frontline worker’ who has to physically travel to work during lockdown? 

Examples include people employed in health and social care, education and childcare, local/national government, food production or sale, the prison service, the police and public transport.

- Yes (1)
- No (0)

| Page Break |
| --- |

Display This Question:

If Q19 = Yes

| 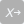 | 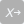 |
| --- | --- |

Q20 Do you work in any of the following places?
Please select all that apply.

- Community healthcare (e.g. doctors’ surgery, dental surgery, health clinic) (0)
- Funeral parlour or mortuary (1)
- Hospital/COVID-19 treatment centre (2)
- Medical laboratory (3)
- Nursing home or hospice (4)
- People’s private homes (5)
- Police station (6)
- Prison (7)
- Residential care home (8)
- School or nursery (9)
- Shop or supermarket (10)
- Transportation vehicle (e.g. bus driver, taxi driver) (11)
- Other (12)

| Page Break |
| --- |

| 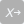 | 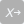 |
| --- | --- |

Q21 What is the highest level of education that you have completed?

- Primary school (0)
- Secondary school up to 16 years (1)
- Higher or secondary or further education (A-levels, BTEC, etc.) (2)
- College or university (3)
- Post-graduate degree (4)

| Page Break |
| --- |

| 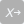 | 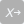 |
| --- | --- |

Q22 Please select the box that best describes your current housing situation:

- I own my home outright (0)
- I own my home and I am paying a mortgage (1)
- I am renting privately (2)
- I am renting from the council/housing association (3)
- I am staying with friends or family (4)
- I am homeless or living in temporary accommodation (5)
- Other (6)

| Page Break |
| --- |

| 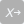 | 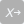 |
| --- | --- |

Q23 How many bedrooms are there in your current accommodation?

▼ 1 (1) ... 10+ (10)

| Page Break |
| --- |

| 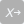 | 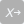 |
| --- | --- |

Q24 Do you live alone?

- Yes (1)
- No (0)

| Page Break |
| --- |

Display This Question:

If Q24 = No

| 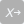 | 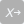 |
| --- | --- |

Q25 How many people other than yourself live in your household?

| Children aged 0-4 years (25) | ▼ 0 (154) ... 10+ (153) |
| --- | --- |
| Children aged 5-15 years (26) | ▼ 0 (154) ... 10+ (153) |
| People aged 16-64 years (27) | ▼ 0 (154) ... 10+ (153) |
| People aged 65 years or more (28) | ▼ 0 (154) ... 10+ (153) |

| Page Break |
| --- |

| 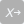 | 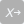 |
| --- | --- |

Q26 Does your household have any pets?

- Yes (1)
- No (0)

| Page Break |
| --- |

Display This Question:

If Q26 = Yes

| 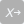 | 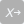 |
| --- | --- |

Q27 Please indicate which types of pet you have at home. 
Select all that apply.

- Cat (0)
- Dog (1)
- Indoor bird (e.g. budgie, parrot, canary) (2)
- Rabbit / Guinea pig / Hamster (3)
- Tortoise, turtle, lizard or snake (4)
- Other (5)

| Page Break |
| --- |

| 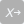 | 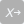 |
| --- | --- |

Q28 Do you have regular contact with any of the following live animals or birds? 
Select all that apply.

- Poultry (Chickens, Ducks, Turkeys, Geese) (0)
- Pigs (1)
- Sheep (2)
- Goats (3)
- Cows (4)
- Horses (5)
- Exotic animals (e.g. alpaca, llama, zoo animals) (6)
- ⊗None of the above (7)

| Page Break |
| --- |

| 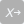 | 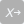 |
| --- | --- |

Q29 In the last month, was your household income sufficient to cover the basic needs of your household, such as food and heating?

- Yes (1)
- Mostly (3)
- Sometimes (2)
- No (0)

| Page Break |
| --- |

Q29.5 How does your household income today compare with how it was in January 2020?

- Higher (1)
- The same (2)
- Lower (3)
- Can't remember / not sure / don't know (4)

| Page Break |
| --- |

| 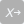 | 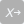 |
| --- | --- |

Q30 Do you currently claim Universal Credit?

- Yes, I have applied to receive Universal Credit but have **not yet** received any payments (1)
- Yes, I have claimed Universal Credit and received **one or more** payments (2)
- No (0)

End of Block: COVIDENCE_Demographics

Start of Block: COVIDENCE_Lifestyle

Q31 The next section asks some questions about your lifestyle and diet.

| 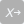 | 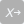 |
| --- | --- |

Q32
Which of these best describes your use of cigarettes?

- I have never smoked cigarettes (0)
- I used to smoke cigarettes occasionally but now not at all (1)
- I used to smoke cigarettes daily but now not at all	(2)
- I smoke cigarettes occasionally but not every day (3)
- I smoke cigarettes daily (4)

| Page Break |
| --- |

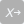

Q33 Which of these best describes your use of e-cigarettes (vaping)?

- I have never vaped or used e-cigarettes (0)
- I used to use e-cigarettes occasionally, but now not at all (1)
- I used to use e-cigarettes daily but now not at all	(2)
- I vape occasionally but not every day (3)
- I vape daily (4)

| Page Break |
| --- |

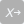

Q34 Are you regularly exposed to smoke from other people’s cigarettes at home or in a car?

- Yes (1)
- No (0)

| Page Break |
| --- |

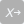

Q35 Roughly how many hours have you spent outdoors in the last week?

▼ 0 (0) ... 10+ (10)

| Page Break |
| --- |

| 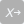 | 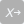 |
| --- | --- |

Q36 During the last week, roughly how many hours did you spend doing light exercise that does not make you particularly breathless, such as light gardening, walking, including walking for pleasure or exercise, walking to the shops, walking to work?

▼ 0 (28) ... 10+ (38)

| Page Break |
| --- |

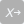

Q37 During the last week, roughly how many hours did you spend doing more vigorous physical exercise of sufficient intensity to make you breathless or to raise your heart rate significantly, such as heavy physical work, more strenuous gardening (e.g. vigorous digging, landscaping) swimming, jogging, aerobics, football, tennis, cycling, gym workout?

▼ 0 (0) ... 10+ (10)

| Page Break |
| --- |

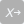

Q38 During the last week, roughly how many hours did you spend doing lower impact physical exercise to improve flexibility or core strength such as yoga, tai chi or pilates?

▼ 0 (0) ... 10+ (10)

| Page Break |
| --- |

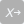

Q39 How many units of alcohol did you drink over the last 7 days? 

One unit is a ½ a pint (285 ml) of ordinary beer, lager or cider; 25ml of spirits; 1 small glass (75ml) of wine; or 50ml of sherry.

- None (0)
- 1-7 units (1)
- 8-14 units (2)
- 15-21 units (3)
- 22-28 units (4)
- More than 28 units (5)

| Page Break |
| --- |

| 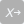 | 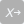 |
| --- | --- |

Q40 During the past month, what time have you usually **gone to bed** in the evening (turn out the lights in order to sleep)?
 
*Please enter time in 24 hour clock (for example, 3:00 is 3:00am while 15:00 is 3:00pm).*

|  | Hour | Minutes |
| --- | --- | --- |
| Time: (NES.18.0) | ▼ 1 (1 ... 24 (24) | ▼ 00 (1 ... 45 (4) |

| Page Break |
| --- |

| 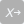 | 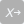 |
| --- | --- |

Q41
During the past month, what time did you usually **get up** in the morning? 

*Please enter time in 24 hour clock (for example, 3:00 is 3:00am while 15:00 is 3:00pm).*

|  | Hour | Minutes |
| --- | --- | --- |
| Time: (NES.18.0) | ▼ 1 (1 ... 24 (24) | ▼ 00 (1 ... 45 (4) |

| Page Break |
| --- |

| 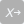 | 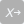 |
| --- | --- |

Q42 During the past month, how many hours of actual sleep did you get per night on average? 
(This may be different than the number of hours you spent in bed)

▼ 0 (1) ... 24 (25)

| Page Break |
| --- |

End of Block: COVIDENCE_Lifestyle

Start of Block: COVIDENCE_Diet

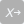

Q43 Do you **exclude** any of the following foods from your diet?

Select all that apply.

- Eggs (0)
- Cow’s milk or products made from cow’s milk (e.g. cheese, yoghurt) (1)
- Fish (2)
- White meat (e.g. poultry) (3)
- Red meat (4)
- ⊗No, I eat all of these foods (5)

| Page Break |
| --- |

Q44 Over the **last week**, how many **portions** of the following did you eat **per day**, on average?

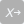

Q45 Fruit, vegetables and salad?

 *1 portion = 80g (e.g. one apple or two broccoli spears or 3 tablespoons of peas or carrots or one bowl of salad)*

▼ 0 (0) ... 10 or more (10)

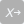

Q46 Starchy foods, e.g. potatoes, bread, rice, pasta 

*1 portion = a slice of bread or a potato about the size of your fist (220g) or two handfuls of dried pasta shapes or rice (75g) or three handfuls of breakfast cereal*

▼ 0 (0) ... 10 or more (10)

| Page Break |
| --- |

Q47 Over the **last week**, how many **portions** of the following did you eat **per day**, on average?

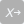

Q48 Protein-rich foods from plant sources, e.g. beans or pulses.

*1 portion = a small (200g) can of baked beans or three heaped tablespoons (200g) of other pulses (e.g. kidney beans, chick peas).*

▼ 0 (0) ... 10 or more (10)

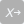

Q49
Protein-rich foods from animal sources, e.g. eggs, fish, chicken, red meat 

*1 portion = two eggs or three fish fingers or a small (160g) tin of tuna or a piece of fish, chicken or red meat about the size of your palm (140g) or two sausages or five slices of ham.*

▼ 0 (0) ... 10 or more (10)

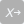

Q50 Dairy products (e.g. cow’s milk, cheese, yoghurts) or calcium-fortified dairy alternatives (e.g. soya milks, soya yoghurts and soya cheeses) 

*1 portion = a cup of milk, a standard pot of yogurt or a piece of cheese about the size of two thumbs together (30g).*

▼ 0 (0) ... 10 or more (10)

| Page Break |
| --- |

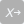

Q51 Over the last week, how many cups or glasses of fluid did you drink per day, on average?

*1 cup or glass = about 150mL. Non-alcoholic drinks including water, tea, coffee, milk and other soft drinks all count.*

▼ 0 (0) ... 10+ (10)

| Page Break |
| --- |

Q52 The next two questions ask about the amount of fish you have eaten in the last week. 
We will ask about oily fish and non-oily fish (white fish and seafood) separately.

| Page Break |
| --- |

Display This Question:

If Q43 != Fish

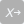

Q53 Over the last week, how many portions of **oily** fish did you eat? 
e.g. herring, pilchards, salmon, sardines, sprats, trout and mackerel. 

*1 portion = a small tin of oily fish (around 100g) or a piece of oily fish about the size of your palm (140g).*

▼ 0 (0) ... 10+ (10)

Display This Question:

If Q43 != Fish

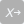

Q54 Over the last week, how many portions of **white fish or seafood (i.e non-oily fish)** did you eat? 
e.g. cod, haddock, plaice, prawns and tuna

*1 portion = a small (160g) tin of tuna or a piece of white fish about the size of your palm (140g) or 100g prawns*

▼ 0 (0) ... 10+ (10)

| Page Break |
| --- |

End of Block: COVIDENCE_Diet

Start of Block: COVIDENCE_Nutritional Supplements

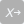

Q55 Over the **last month**, have you taken any of the following supplements at least **once per week**? 
Select all that apply.

- Multivitamin (including prenatal multivitamins) (0)
- Supplement containing vitamin A only (1)
- Supplement containing vitamin B only (2)
- Supplement containing vitamin C only (3)
- Supplement containing vitamin D only (4)
- Supplement containing calcium only (5)
- Supplement containing calcium and vitamin D combined (6)
- Supplement containing vitamin E only (7)
- Supplement containing zinc only (8)
- Supplement containing iron only (9)
- Supplement containing probiotics (10)
- Supplement containing fish oil, krill oil or other source of omega-3 fatty acids (11)
- Supplement containing cod liver oil (12)
- Supplement containing echinacea (13)
- Supplement containing garlic or garlic powder (allicin) (14)
- Supplement containing turmeric / curcumin (15)
- Supplement containing Cannabidiol (CBD) oil (16)
- Supplements containing folic acid (17)
- Supplement containing Selenium only (18)
- Other (e.g. other micronutrients (such as herbal supplements) or combinations of micronutrients (such vitamin C & zinc)) Please specify: (19) ________________________________________________
- ⊗None of the above (20)

| Page Break |
| --- |

Display This Question:

If Q55 = Multivitamin (including prenatal multivitamins)

Or Q55 = Supplement containing vitamin D only

Or Q55 = Supplement containing calcium and vitamin D combined

Or Q55 = Other (e.g. other micronutrients (such as herbal supplements) or combinations of micronutrients (such vitamin C & zinc)) Please specify:

Q55.1_ud   You have indicated that you take one or more nutritional supplements that may contain vitamin D. Please indicate the amount of vitamin D that is in each dose you take (if any). 

- None (1)
- 1 to 200 IU (0.1 to 5 micrograms) (14)
- 201 to 400 IU (5.1 to 10 micrograms) (2)
- 401 to 800 IU (10.1 to 20 micrograms) (3)
- 801 to 3,200 IU (20.1 to 80 micrograms) (4)
- More than 3,200 IU (80 micrograms) (5)
- Don't know / not sure (6)

| Page Break |
| --- |

Display This Question:

If Q55 = Multivitamin (including prenatal multivitamins)

Or Q55 = Supplement containing vitamin D only

Or Q55 = Supplement containing calcium and vitamin D combined

Or Q55 = Other (e.g. other micronutrients (such as herbal supplements) or combinations of micronutrients (such vitamin C & zinc)) Please specify:

Q55.2_ud Now, please tell us how often you take this dose.

- Every day (1)
- 2-6 times a week (2)
- Once a week (3)
- 2-3 times a month (4)
- Once a month (5)
- Less often than once a month (6)
- Don't know / not sure (7)

| Page Break |
| --- |

Display This Question:

If Q55 = Multivitamin (including prenatal multivitamins)

Or Q55 = Supplement containing vitamin D only

Or Q55 = Supplement containing calcium and vitamin D combined

Or Q55 = Other (e.g. other micronutrients (such as herbal supplements) or combinations of micronutrients (such vitamin C & zinc)) Please specify:

Q55.1_ud.1 If there is additional information about your vitamin D intake that our questions haven’t captured, please write it here (optional)

________________________________________________________________

End of Block: COVIDENCE_Nutritional Supplements

Start of Block: COVIDENCE_General Health

Q56 The following questions will ask about your general health.

| Page Break |
| --- |

Display This Question:

If sex = Female

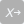

Q57 Are you currently pregnant?

- Yes (1)
- No (0)

| Page Break |
| --- |

Display This Question:

If Q57 = Yes

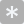

Q58 What is your estimated date of delivery? (DD/MM/YYYY)
e.g. 25/04/2020

________________________________________________________________

| Page Break |
| --- |

Display This Question:

If Q57 = Yes

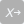

Q59 Is this pregnancy a multiple pregnancy? (e.g. twins/triplets/etc.)

- Yes (1)
- No (0)

| Page Break |
| --- |

Display This Question:

If Q57 = Yes

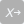

Q60 Have you experienced any of the following complications during this pregnancy?

- Pre-eclampsia/high blood pressure (0)
- Gestational diabetes (1)
- Other complication (2) ________________________________________________
- ⊗None of the above (3)

| Page Break |
| --- |

Display This Question:

If Q57 = Yes

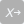

Q61 How many pregnancies of 24 weeks or longer have you had, including this one?

▼ 0 (0) ... 10+ (10)

| Page Break |
| --- |

Display This Question:

If Q57 = Yes

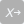

Q62 How many pregnancies of less than 24 weeks have you had, including this one?

▼ 0 (0) ... 10+ (10)

| Page Break |
| --- |

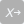

Q63 Over the last 12 months, would you say that on the whole, your health has been:

- Excellent	(0)
- Very good (1)
- Good (2)
- Fair (3)
- Poor (4)

End of Block: COVIDENCE_General Health

Start of Block: COVIDENCE_Quality of Life

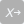

Q64 Which of the following statements best describes your breathing currently?

- I get breathless only with strenuous exercise (0)
- I get short of breath when hurrying on level ground or up a slight hill (1)
- I am slower than most people of the same age on a level surface or have to stop when walking at my own pace on the level (2)
- I stop for breath walking 100 metres or after a walking few minutes at my own pace on the level (3)
- I am too breathless to leave the house (4)

| Page Break |
| --- |

Q64.a We would now like to ask three questions about symptoms that have been reported by people who are recovering from COVID-19. We would like to assess all COVIDENCE UK participants for these symptoms, so that we can compare them in people who have and haven’t had COVID-19.

| Page Break |
| --- |

Q64.1_ud First, have you or your family noticed that you are currently experiencing any of the following problems? 
Please answer all of these questions **WHETHER OR NOT YOU HAVE HAD DEFINITE OR SUSPECTED COVID-19**.

|  | No (1) | Yes but improving (2) | Yes, but not improving or worsening (3) | Yes and worsening (4) |
| --- | --- | --- | --- | --- |
| Excessive shortness of breath (1) |  |  |  |  |
| Coughing (2) |  |  |  |  |
| Unusual tiredness or fatigue (3) |  |  |  |  |
| Problems with sleeping (4) |  |  |  |  |
| Memory problems (5) |  |  |  |  |
| Difficulty concentrating (6) |  |  |  |  |
| Pains in your muscles or joints (7) |  |  |  |  |
| Problems with your sense of smell / taste (8) |  |  |  |  |
| Diarrhoea (9) |  |  |  |  |
| Stomach (abdominal) pains (10) |  |  |  |  |
| Changes to your voice (11) |  |  |  |  |
| Hair loss (12) |  |  |  |  |
| Unusual racing of the heart (13) |  |  |  |  |
| Lightheadedness or dizziness (14) |  |  |  |  |
| Unusual sweating (15) |  |  |  |  |

| Page Break |
| --- |

Q64.2_ud
Second, here is a list of statements relating to common long-term symptoms of COVID-19. Please select one answer per line to indicate how you have been feeling over the past 7 days. 

Please answer all of these questions **WHETHER OR NOT YOU HAVE HAD DEFINITE OR SUSPECTED COVID-19**.

|  | Not at all (1) | A little bit (2) | Somewhat (3) | Quite a bit (4) | Very much (5) |
| --- | --- | --- | --- | --- | --- |
| I feel fatigued (1) |  |  |  |  |  |
| I feel weak all over (2) |  |  |  |  |  |
| I feel listless ("washed out") (3) |  |  |  |  |  |
| I feel tired (4) |  |  |  |  |  |
| I have trouble starting things because I am tired (5) |  |  |  |  |  |
| I have trouble finishing things because I am tired (6) |  |  |  |  |  |
| I have energy (7) |  |  |  |  |  |
| I am able to do my usual activities (8) |  |  |  |  |  |
| I need to sleep during the day (9) |  |  |  |  |  |
| I am too tired to eat (10) |  |  |  |  |  |
| I need help doing my usual activities (11) |  |  |  |  |  |
| I am frustrated by being too tired to do the things I want to do (12) |  |  |  |  |  |
| I have to limit my social activity because I am tired (13) |  |  |  |  |  |

| Page Break |
| --- |

Q64.3_ud Third, would YOU say that you currently have 'long COVID', i.e. ongoing symptoms more than four weeks after the onset of proven or suspected COVID-19?

- Yes (1)
- No (2)
- Don't know / not sure (3)

| Page Break |
| --- |

Q65 Under each heading, please click the ONE box that best describes your health TODAY.

| 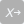 | 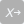 |
| --- | --- |

Q66 Mobility

- I have no problems in walking about (0)
- I have some problems in walking about (1)
- I am confined to bed (2)

| Page Break |
| --- |

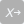

Q67 Self-Care

- I have no problems with self-care (0)
- I have some problems washing or dressing myself (1)
- I am unable to wash or dress myself (2)

| Page Break |
| --- |

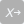

Q68 Usual Activities (e.g. work, study, housework, family or leisure activities)

- I have no problems with performing my usual activities (0)
- I have some problems with performing my usual activities (1)
- I am unable to perform my usual activities (2)

| Page Break |
| --- |

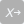

Q69 Pain / Discomfort

- I have no pain or discomfort (0)
- I have moderate pain or discomfort (1)
- I have extreme pain or discomfort (2)

| Page Break |
| --- |

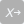

Q70 Anxiety / Depression

- I am not anxious or depressed (0)
- I am moderately anxious or depressed (1)
- I am extremely anxious or depressed (2)

End of Block: COVIDENCE_Quality of Life

Start of Block: COVIDENCE_Your own health state today

Display This Question:

If sex = Male

And sex = Female

Q71
We would like to know how good or bad your health is TODAY. 
This scale is numbered from 0 to 100. 
100 means the best health you can imagine. 
0 means the worst health you can imagine.  
Please click on the scale to indicate how your health is TODAY.

|  | The worst health you can imagine | The best health you can imagine |
| --- | --- | --- |

|  | 0 | 10 | 20 | 30 | 40 | 50 | 60 | 70 | 80 | 90 | 100 |
| --- | --- | --- | --- | --- | --- | --- | --- | --- | --- | --- | --- |

| Please click on the scale to indicate how your health is TODAY. () | 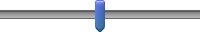 |
| --- | --- |

End of Block: COVIDENCE_Your own health state today

Start of Block: COVIDENCE_Your own health state today_updated

| 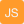 | 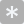 |
| --- | --- |

Q71_updated
 We would like to know how good or bad your health is TODAY. 
 This scale is numbered from 0 to 100. 	100 means the best health you can imagine. 	0 means the worst health you can imagine.  
 Please click on the scale to indicate how your health is TODAY.
 The best health you can imagine
 The worst health you can imagine

________________________________________________________________

End of Block: COVIDENCE_Your own health state today_updated

Start of Block: COVIDENCE_Recent Health

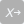

Q72 Since February 1st 2020, have you experienced any of the following symptoms: loss of smell or taste, fever, persistent cough, fatigue, diarrhoea, abdominal pain or loss of appetite?

- Yes, I have had one or more of these symptoms since 1st of February (0)
- No, I have not had any of these symptoms since 1st of February (1)

Skip To: Q102 If Q72 = No, I have not had any of these symptoms since 1st of February

| Page Break |
| --- |

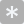

Q73 When did your symptoms start? (DD/MM/YYYY)
*e.g. 25/04/2020*

________________________________________________________________

| Page Break |
| --- |

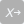

Q74 How ill did you feel at your worst?

- Mildly unwell - I could do most of my usual activities (0)
- Moderately unwell - I couldn’t do usual activities, but didn’t need to go to bed in the daytime (1)
- Very unwell – I had to go to bed in the daytime (2)

| Page Break |
| --- |

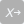

Q75 Did you have a fever (high temperature)?

- Yes (1)
- No (0)

| Page Break |
| --- |

Display This Question:

If Q75 = Yes

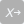

Q76 What was your highest temperature?

- I don’t know / I did not take my temperature (0)
- Less than 38°C (less than 100°F) (1)
- 38.0-39.0°C (100-102°F) (2)
- More than 39°C (more than 102°F) (3)

| Page Break |
| --- |

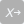

Q77 Did you have a persistent cough (coughing a lot for more than an hour, or 3 or more coughing episodes in 24 hours)?

- No (0)
- Persistent dry cough (i.e. producing little or no phlegm) (1)
- Persistent productive cough (2)

| Page Break |
| --- |

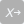

Q78 Did you experience unusual fatigue?

- No (0)
- Mild fatigue (1)
- Severe fatigue - I struggled to get out of bed (2)

| Page Break |
| --- |

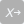

Q79 Did you have a headache?

- Yes (1)
- No (0)

| Page Break |
| --- |

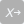

Q80 Did you have muscle aches?

- Yes (1)
- No (0)

| Page Break |
| --- |

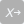

Q81 Did you have a loss of sense of smell or taste?

- Yes (1)
- No (0)

| Page Break |
| --- |

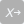

Q82  Did you experience unusual shortness of breath?

- No (0)
- Yes, mild symptoms - slight shortness of breath during ordinary activity (1)
- Yes, significant symptoms – breathing was comfortable only at rest (2)
- Yes, severe symptoms - breathing was difficult even at rest (3)

| Page Break |
| --- |

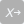

Q83 Please indicate any other symptoms you had. 
Select as many that apply:

- Sore throat (0)
- Sneezing (1)
- Runny nose (2)
- Blocked nose (3)
- Unusually hoarse voice (4)
- Unusual Chest pain (5)
- Unusual Abdominal pain (6)
- Diarrhoea (7)
- Confusion, disorientation, or drowsiness (8)
- Other symptom (please specify) (9) ________________________________________________
- ⊗None of the above (10)

| Page Break |
| --- |

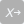

Q84 Did you skip any meals because you felt unwell?

- Yes (1)
- No (0)

| Page Break |
| --- |

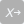

Q85 Did you take any over the counter medication to relieve your symptoms?

- Yes (1)
- No (0)

Display This Question:

If Q85 = Yes

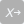

Q86 You stated that you took over-the-counter medication, please click as many as apply:

- Paracetamol tablets (0)
- Cold and flu relief tablets or medicine (1)
- Ibuprofen (2)
- Naproxen (3)
- Aspirin (4)
- Other (5)

| Page Break |
| --- |

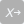

Q87 Were you admitted to hospital for treatment of these symptoms? 

- Yes (1)
- No (0)

Skip To: Q94 If Q87 = No

| Page Break |
| --- |

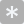

Q88 On what date were you admitted to hospital? (DD/MM/YYYY)

*Note that the year must be typed in full, e.g. 25/04/2020*

________________________________________________________________

| Page Break |
| --- |

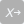

Q89 What treatment did you receive while in the hospital? 
Please select as many that apply:

- Oxygen and fluids (breathing support administered through an oxygen mask, no pressure applied) (0)
- Non-invasive ventilation (breathing support administered through an oxygen mask, which pushes oxygen into your lungs) (1)
- Invasive ventilation (breathing support administered through an inserted tube, people are usually asleep during this procedure) (2)
- I'm not sure (3)
- ⊗None (4)

| Page Break |
| --- |

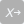

Q90 Have you been discharged from hospital?

- Yes (1)
- No (0)

| Page Break |
| --- |

Display This Question:

If Q90 = Yes

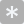

Q91 On what date were you discharged from hospital? (DD/MM/YYYY)

*Note that the year must be typed in full, e.g. 25/04/2020*

________________________________________________________________

| Page Break |
| --- |

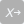

Q92 What did the hospital doctors diagnose?

- Suspected or proven COVID-19 (0)
- Something else (1)

| Page Break |
| --- |

Display This Question:

If Q92 = Something else

Q93 Please write the diagnosis here:

________________________________________________________________

| Page Break |
| --- |

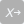

Q94 Do you feel back to normal now?

- Yes (1)
- No (0)

| Page Break |
| --- |

Display This Question:

If Q94 = Yes

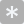

Q95 On what date did you first feel back to normal? (DD/MM/YYYY)

*Note that the year must be typed in full, e.g. 25/04/2020*

________________________________________________________________

| Page Break |
| --- |

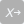

Q96 Since February 1st 2020, have you had a nose/throat swab to test for COVID-19?

- Yes (1)
- No (0)

| Page Break |
| --- |

Display This Question:

If Q96 = Yes

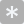

Q97 On what date did you have this nose/throat swab? 
If you are not sure of the exact date, enter the approximate date (DD/MM/YYYY). 
*e.g. 25/04/2020*

________________________________________________________________

Display This Question:

If Q96 = Yes

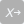

Q98 What was the result?

- Positive (0)
- Negative (1)
- Not known (2)

| Page Break |
| --- |

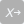

Q99 Since February 1st 2020, have you had a blood test (antibody test) for COVID-19?

- Yes (1)
- No (0)

| Page Break |
| --- |

Display This Question:

If Q99 = Yes

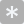

Q100 On what date did you have this blood test (antibody test)? 
If you are not sure of the exact date, enter the approximate date (DD/MM/YYYY).
*e.g. 25/04/2020*

________________________________________________________________

Display This Question:

If Q99 = Yes

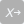

Q101 What was the result?

- Positive (0)
- Negative (1)
- Not known (2)

| Page Break |
| --- |

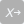

Q102 **Since 1st February 2020,** has any member of your household OTHER THAN YOURSELF developed any of the following symptoms: cough, fever, unusual shortness of breath, unusual fatigue, loss of sense of taste or smell, chest pain, hoarse voice, abdominal pain or diarrhoea?

- Yes (1)
- No (0)
- Not applicable (e.g. I live on my own) (2)

| Page Break |
| --- |

Display This Question:

If Q102 = Yes

Q103 Please provide details of their age and sex, the date when their symptoms first occurred, and results of any testing done for COVID-19. Use one row per person.
*Note that the year must be typed in full, e.g. 25/04/2020*

|  | Age | Sex | | COVID-19 nose/throat swab result | Date when first symptom occurred |
| --- | --- | --- | --- | --- | --- |
|  | (in years) (1) | Male (1) | Female (2) |  | DD/MM/YYYY e.g. 25/04/2020 (1) |
| Person 1 (1) |  |  |  | ▼ Positive (1 ... Not done (4) |  |
| Person 2 (2) |  |  |  | ▼ Positive (1 ... Not done (4) |  |
| Person 3 (4) |  |  |  | ▼ Positive (1 ... Not done (4) |  |
| Person 4 (5) |  |  |  | ▼ Positive (1 ... Not done (4) |  |
| Person 5 (6) |  |  |  | ▼ Positive (1 ... Not done (4) |  |
| Person 6 (12) |  |  |  | ▼ Positive (1 ... Not done (4) |  |
| Person 7 (13) |  |  |  | ▼ Positive (1 ... Not done (4) |  |
| Person 8 (14) |  |  |  | ▼ Positive (1 ... Not done (4) |  |
| Person 9 (15) |  |  |  | ▼ Positive (1 ... Not done (4) |  |
| Person 10 (16) |  |  |  | ▼ Positive (1 ... Not done (4) |  |
| Person 11 (17) |  |  |  | ▼ Positive (1 ... Not done (4) |  |
| Person 12 (18) |  |  |  | ▼ Positive (1 ... Not done (4) |  |
| Person 13 (19) |  |  |  | ▼ Positive (1 ... Not done (4) |  |
| Person 14 (20) |  |  |  | ▼ Positive (1 ... Not done (4) |  |
| Person 15 (21) |  |  |  | ▼ Positive (1 ... Not done (4) |  |
| Person 16 (23) |  |  |  | ▼ Positive (1 ... Not done (4) |  |
| Person 17 (24) |  |  |  | ▼ Positive (1 ... Not done (4) |  |
| Person 18 (25) |  |  |  | ▼ Positive (1 ... Not done (4) |  |
| Person 19 (26) |  |  |  | ▼ Positive (1 ... Not done (4) |  |
| Person 20 (27) |  |  |  | ▼ Positive (1 ... Not done (4) |  |

End of Block: COVIDENCE_Recent Health

Start of Block: COVIDENCE_Long-standing Conditions

| 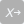 | 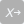 |
| --- | --- |

Q104
Have you been advised by a doctor or other professional that you should be ‘SHIELDED’ during the coronavirus outbreak? ‘Shielding’ involves staying at home and minimising face-to-face contact with people outside the home.

This advice has been given to people with certain underlying conditions that place them at increased risk of severe illness from COVID-19, including solid organ transplant recipients; people with specific cancers; people with severe respiratory conditions; people with severe combined immunodeficiency (SCID) or homozygous sickle cell disease; people on certain immunosuppressive treatments; and pregnant women with significant heart disease. 

- Yes (1)
- No (0)

| Page Break |
| --- |

Display This Question:

If Q104 = Yes

Q105 Please write the name of the underlying condition for which you are being shielded here:

________________________________________________________________

| Page Break |
| --- |

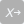

Q106_ud Have you ever been diagnosed with any of the following conditions by a doctor? 

Select all that apply

- Asthma (1)
- Atopic Eczema or Atopic Dermatitis (2)
- Autoimmune disease (e.g. rheumatoid arthritis, multiple sclerosis (MS), lupus (SLE), Crohn’s disease, ulcerative colitis, psoriasis, Raynaud’s disease, scleroderma) (3)
- Cancer (4)
- Cerebral Palsy (5)
- COPD (including chronic bronchitis, and emphysema) (6)
- Cystic Fibrosis (7)
- Dementia (8)
- Diabetes or pre-diabetes (9)
- Hayfever or Allergic Rhinitis (10)
- Heart Attack, Angina or Coronary Artery Disease (11)
- Heart Failure (12)
- High Blood Pressure (Hypertension) (13)
- HIV Infection (14)
- Hyperparathyroidism (overactive parathyroid gland) (15)
- Kidney stones (29)
- Other kidney disease (16)
- Leg Artery Disease (also known as ‘peripheral vascular disease’, ‘peripheral arterial disease’ or ‘intermittent claudication’) (17)
- Mental health disorder (18)
- Motor Neurone Disease (19)
- Organ transplant (20)
- Parkinson's Disease (21)
- Primary immune deficiency (e.g. antibody deficiency, combined immunodeficiency) (22)
- Sarcoidosis (23)
- Sickle Cell Disease (i.e. two copies of altered gene, affected by anaemia and other complications (24)
- Sickle Cell Carrier (also known as ‘sickle cell trait’, with only one copy of altered gene: few symptoms if any) (25)
- Splenectomy (removal of spleen) (26)
- Stroke or Mini-Stroke (27)
- Tuberculosis (TB) (28)
- ⊗None of the above (0)

Skip To: Q136 If Q106_ud = None of the above

Skip To: Q136 If Condition: Have you ever been diagnose... Is Equal to 0. Skip To: Do you have any other long-standing h....

| Page Break |
| --- |

Display This Question:

If Q106_ud != None of the above

And And Have you ever been diagnosed with any of the following conditions by a doctor?  Select all that a... q://QID56/SelectedChoicesCount Is Greater Than 0

| 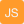 | 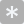 | 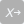 |
| --- | --- | --- |

Q107 Please indicate below your **age at diagnosis** for the following conditions you have been diagnosed with. If you have more than one condition in a given category, write down your age when the first of these was diagnosed.

|  | Age at diagnosis |
| --- | --- |
|  | Age (1) |
| Asthma (Q106_0) |  |
| Atopic Eczema or Atopic Dermatitis (Q106_1) |  |
| Autoimmune disease (e.g. rheumatoid arthritis, multiple sclerosis (MS), lupus (SLE), Crohn’s disease, ulcerative colitis, psoriasis, Raynaud’s disease, scleroderma) (Q106_2) |  |
| Cancer (Q106_3) |  |
| Cerebral Palsy (Q106_4) |  |
| COPD (including chronic bronchitis, and emphysema) (Q106_5) |  |
| Cystic Fibrosis (Q106_6) |  |
| Dementia (Q106_7) |  |
| Diabetes or pre-diabetes (Q106_8) |  |
| Hayfever or Allergic Rhinitis (Q106_9) |  |
| Heart Attack, Angina or Coronary Artery Disease (Q106_10) |  |
| Heart Failure (Q106_11) |  |
| High Blood Pressure (Hypertension) (Q106_12) |  |
| HIV Infection (Q106_13) |  |
| Hyperparathyroidism (Q106_14) |  |
| Kidney disease (Q106_15) |  |
| Leg Artery Disease (Q106_16) |  |
| Motor Neurone Disease (Q106_17) |  |
| Organ transplant (Q106_18) |  |
| Parkinson's Disease (Q106_19) |  |
| Primary immune deficiency (Q106_20) |  |
| Sarcoidosis (Q106_21) |  |
| Sickle Cell Disease (Q106_22) |  |
| Sickle Cell Carrier (Q106_23) |  |
| Splenectomy (Removal of Spleen) (Q106_24) |  |
| Stroke or Mini-Stroke (Q106_25) |  |
| Tuberculosis (TB) (Q106_26) |  |

| Page Break |
| --- |

Display This Question:

If Q106_ud = Asthma

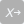

Q108 If you have been diagnosed with Asthma, click as many as apply:

- I sometimes use a reliever inhaler (e.g. ventolin, salbutamol) to control my asthma symptoms (0)
- I take a regular inhaler that includes a steroid preventer ONLY (e.g. Beclomethasone, Budesonide, Ciclesonide, Fluticasone or Mometasone) (1)
- I take a regular inhaler that includes a long-acting bronchodilator ONLY (e.g. salmeterol, formoterol) (2)
- I take a regular combination inhaler that contains BOTH a steroid preventer AND a long-acting bronchodilator (e.g. Seretide, Symbicort, Flutiform, Fostair) (3)
- My asthma is being treated with monoclonal antibody infusions at the hospital (4)
- In the last 12 months I have had one or more asthma attacks requiring treatment with steroid tablets (prednisolone) (5)
- In the last 12 months I have had one or more asthma attacks requiring hospital admission (6)
- ⊗None of the above (7)

| Page Break |
| --- |

Display This Question:

If Q106_ud = Primary immune deficiency (e.g. antibody deficiency, combined immunodeficiency)

Q109 You indicated that you have a primary immune deficiency. Please type the name of your condition in the box below:

________________________________________________________________

| Page Break |
| --- |

Display This Question:

If Q106_ud = COPD (including chronic bronchitis, and emphysema)

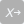

Q110 If you have been diagnosed with COPD (including chronic bronchitis, and emphysema), click as many as apply:

- I sometimes use a reliever inhaler (e.g. ventolin, salbutamol, atrovent, ipratropium) to control my COPD symptoms (0)
- I take a regular inhaler that includes a steroid preventer ONLY (e.g. Beclomethasone, Budesonide, Ciclesonide, Fluticasone or Mometasone) (1)
- I take a regular inhaler that includes a long-acting bronchodilator ONLY (e.g. tiotropium, glycopyronium, aclidinium, salmeterol, formoterol, indacaterol) (2)
- I take a regular combination inhaler that contains BOTH a steroid preventer AND a long-acting bronchodilator (e.g. Duaklir Genuair, Ultbrio Breezhaler, Spiolto Respimat, Anoro Ellipta, Seretide, Symbicort, Flutiform, Fostair) (3)
- In the last 12 months I have had one or more COPD attacks / exacerbations requiring treatment with steroid tables (prednisolone) and / or antibiotics (4)
- In the last 12 months I have had one or more COPD attacks / exacerbations requiring hospital admission (5)
- I am on long term oxygen therapy (i.e. I use an oxygen cylinder at home) (6)
- ⊗None of the above (7)

| Page Break |
| --- |

Display This Question:

If Q106_ud = Heart Attack, Angina or Coronary Artery Disease

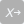

Q111 If you have been diagnosed with Heart Attack, Angina or Coronary Artery Disease, click as many as apply:

- I have had a heart attack diagnosed by a doctor in the past (0)
- I have had a coronary artery bypass graft operation in the past (1)
- I have had a coronary angioplasty and/or coronary artery stent in the past (2)
- I am awaiting hospital admission for coronary angioplasty and/or coronary artery stent (3)
- In the last 12 months I have been admitted to hospital for treatment of chest pain, angina or heart attack (4)
- I have had a coronary angiogram in the past (X-ray picture of the heart arteries following injection of dye via wrist or groin) (5)
- ⊗None of the above (6)

| Page Break |
| --- |

Display This Question:

If Q111 = I have had a coronary angiogram in the past (X-ray picture of the heart arteries following injection of dye via wrist or groin)

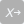

Q112 What did your doctor tell you about your latest angiogram result? Click one option only:

- Normal (0)
- Only one coronary artery affected (1)
- Two coronary arteries affected (2)
- Three coronary arteries affected (3)
- I don’t know the result (4)

| Page Break |
| --- |

Display This Question:

If Q106_ud = Heart Failure

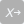

Q113 In the last 12 months have you been admitted to hospital for treatment of heart failure?

- Yes (1)
- No (0)

| Page Break |
| --- |

Display This Question:

If Q106_ud = Heart Failure

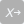

Q114 Which one of the following best describes your symptoms of Heart Failure (select one):

- I have **no symptoms** and **no limitation** in ordinary physical activity, e.g. shortness of breath when walking, climbing stairs etc. (0)
- I have **mild** shortness of breath and/or angina and **slight limitation** during ordinary activity. (1)
- I have **marked limitation** in activity due to my symptoms, even when walking short distances (20—100 m). I am only comfortable when at rest. (2)
- I experience symptoms even while at rest; I am mostly **bedbound**. (3)

| Page Break |
| --- |

Display This Question:

If Q106_ud = Leg Artery Disease (also known as ‘peripheral vascular disease’, ‘peripheral arterial disease’ or ‘intermittent claudication’)

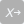

Q115 If you have been diagnosed with Leg Artery Disease (also known as ‘peripheral vascular disease’, ‘peripheral arterial disease’ or ‘intermittent claudication’), click as many as apply:

- I get tingling sensations in my legs (0)
- I get pains or cramps in my calves when I walk, which get better when I rest (1)
- I sometimes get pain in my calves when I am standing still or sitting (2)
- I have ulcers and / or gangrene on my lower legs and/or my feet (3)
- ⊗None of the above (4)

| Page Break |
| --- |

Display This Question:

If Q106_ud = Stroke or Mini-Stroke

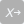

Q116 If you have been diagnosed with Stroke or Mini-Stroke, select one option below which best describes you:

- I have no symptoms at all (0)
- I have no significant disability despite symptoms: able to carry out all usual duties and activities (1)
- I have a slight disability: unable to carry out all previous activities but able to look after own affairs without assistance (2)
- I have a moderate disability: requiring some help, but able to walk without assistance (3)
- I have a moderately severe disability: unable to walk without assistance, and unable to attend to own bodily needs without assistance (4)
- I have a severe disability: bedridden, incontinent, and requiring constant nursing care and attention (5)

| Page Break |
| --- |

Display This Question:

If Q106_ud = Other kidney disease

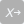

Q117_ud If you have been diagnosed with kidney disease other than kidney stones, click as many as apply:

- I have regular dialysis for kidney failure (0)
- In the last 12 months I have been admitted to hospital for treatment of kidney failure (1)
- ⊗None of the above (2)

| Page Break |
| --- |

Display This Question:

If Q106_ud = High Blood Pressure (Hypertension)

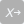

Q118 If you have been diagnosed with High Blood Pressure (Hypertension), click as many as apply:

- I take tablets to treat my high blood pressure (please provide details in medication section below) (0)
- I have kidney problems related to my high blood pressure (1)
- I have eye problems related to my high blood pressure (2)
- In the last 12 months I have been admitted to hospital for treatment of my high blood pressure (3)
- ⊗None of the above (4)

| Page Break |
| --- |

Display This Question:

If Q106_ud = Cancer

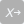

Q119 Do you currently have cancer?

- No, cancer cured or in remission (0)
- Yes, currently receiving treatment (1)

| Page Break |
| --- |

Display This Question:

If Q106_ud = Cancer

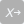

Q120 Please click all cancer sites that apply:

- Bladder (0)
- Bowel (1)
- Brain (2)
- Breast (3)
- Cervix (4)
- Head and Neck (5)
- Kidney (6)
- Leukaemia (7)
- Liver (8)
- Lung (9)
- Lymphoma (10)
- Melanoma Skin Cancer (11)
- Myeloma (12)
- Oesophagus (13)
- Pancreas (14)
- Prostate (15)
- Stomach (16)
- Thyroid (17)
- Uterus (18)
- Other site (19)

| Page Break |
| --- |

Display This Question:

If Q119 = Yes, currently receiving treatment

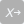

Q121 Please select all treatments that you are currently receiving. 
If you select any below, please provide details in the medication section later on.

Select all that apply:

- I am currently receiving chemotherapy (0)
- I am currently receiving immunotherapy (1)
- I am currently receiving radiotherapy (2)
- I am currently receiving palliative care (symptom control only) (3)
- ⊗None of the above (4)

| Page Break |
| --- |

Display This Question:

If Q106_ud = Diabetes or pre-diabetes

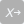

Q122 You indicated you have been diagnosed with diabetes or pre-diabetes. Please specify your diagnosis:

- Pre-diabetes (high blood sugar levels, not enough to be diagnosed with diabetes) (0)
- Type 1 diabetes (1)
- Type 2 diabetes (2)
- Other type of diabetes (3)

| Page Break |
| --- |

Display This Question:

If Q122 = Type 1 diabetes

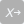

Q123 You indicated Type 1 diabetes, please select as many as apply:

- I use pump therapy (0)
- I use a continuous glucose sensor or flash glucose monitor (1)
- I have had a hospital admission in the last 1 year related to diabetes (2)
- I have had diabetic ketoacidosis before (3)
- I have nerve damage (neuropathy) related to my diabetes (4)
- I have kidney damage (nephropathy) related to my diabetes (5)
- I have foot problems related to my diabetes (6)

sex = Male

- I have trouble getting or keeping an erection as a result of my diabetes (7)
- None of the above (8)

| Page Break |
| --- |

Display This Question:

If Q122 = Type 1 diabetes

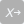

Q124 My last HbA1c (diabetes blood test) was:

- Less than 48mmol/l (6.5%) (0)
- 48-57mmol/l (6.5-7.4%) (1)
- 58-69mmol/l (7.5-8.4%) (2)
- 70-79mmol/l (8.5-9.4%) (3)
- 80 or above (9.5%) (4)
- Not known (5)

| Page Break |
| --- |

Display This Question:

If Q122 = Type 2 diabetes

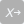

Q125 You indicated Type 2 diabetes, please select as many as apply:

- I have had a hospital admission in the last 1 year related to diabetes (0)
- I have nerve damage (neuropathy) related to my diabetes (1)
- I have kidney damage (nephropathy) related to my diabetes (2)
- I have foot problems related to my diabetes (3)

sex = Male

- I have trouble getting or keeping an erection as a result of my diabetes (4)
- None of the above (5)

| Page Break |
| --- |

Display This Question:

If Q122 = Type 2 diabetes

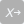

Q126 My last HbA1c (diabetes blood test) was:

- Less than 48mmol/l (6.5%) (0)
- 48-57mmol/l (6.5-7.4%) (1)
- 58-69mmol/l (7.5-8.4%) (2)
- 70-79mmol/l (8.5-9.4%) (3)
- 80 or above (9.5%) (4)
- Not known (5)

| Page Break |
| --- |

Display This Question:

If Q122 = Other type of diabetes

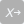

Q127 You indicated other type of diabetes, please select as many as apply:

- I have had a hospital admission in the last 1 year related to diabetes (0)
- I have nerve damage (neuropathy) related to my diabetes (1)
- I have kidney damage (nephropathy) related to my diabetes (2)
- I have foot problems related to my diabetes (3)

sex = Male

- I have trouble getting or keeping an erection as a result of my diabetes (4)
- None of the above (5)

| Page Break |
| --- |

Display This Question:

If Q122 = Other type of diabetes

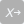

Q128 My last HbA1c (diabetes blood test) was:

- Less than 48mmol/l (6.5%) (0)
- 48-57mmol/l (6.5-7.4%) (1)
- 58-69mmol/l (7.5-8.4%) (2)
- 70-79mmol/l (8.5-9.4%) (3)
- 80 or above (9.5%) (4)
- Not known (5)

| Page Break |
| --- |

Display This Question:

If Q106_ud = Organ transplant

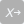

Q129 You indicated you have had an organ transplant, please select as many as apply:

- Kidney transplant (0)
- Heart transplant (1)
- Liver transplant (2)
- Lung transplant (3)
- Bone marrow transplant (4)
- Other transplant (5)

| Page Break |
| --- |

Display This Question:

If Q106_ud = Autoimmune disease (e.g. rheumatoid arthritis, multiple sclerosis (MS), lupus (SLE), Crohn’s disease, ulcerative colitis, psoriasis, Raynaud’s disease, scleroderma)

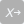

Q130 You indicated you have been diagnosed with an autoimmune disease (e.g. rheumatoid arthritis, multiple sclerosis (MS), lupus (SLE), Crohn’s disease, ulcerative colitis, psoriasis). 

Please specify your diagnosis or diagnoses by clicking as many as apply:

- Autoimmune thyroid disease (0)
- Crohn’s disease (1)
- Lupus (SLE) (2)
- Multiple sclerosis (MS) (3)
- Psoriasis (4)
- Reynaud’s disease (5)
- Rheumatoid arthritis (6)
- Scleroderma (7)
- Type 1 diabetes (8)
- Ulcerative colitis (9)
- Other (10)

| Page Break |
| --- |

Display This Question:

If Q106_ud = Organ transplant

Or Q106_ud = Autoimmune disease (e.g. rheumatoid arthritis, multiple sclerosis (MS), lupus (SLE), Crohn’s disease, ulcerative colitis, psoriasis, Raynaud’s disease, scleroderma)

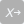

Q131 Are you taking medicines to suppress the immune system? 
(Please provide details in the medication section towards the end of the questionnaire)

- Yes (1)
- No (0)

| Page Break |
| --- |

Q131.1 We would now like to ask about your dental health, as evidence is emerging that this may affect susceptibility to COVID-19.

Gum disease (periodontal disease, periodontitis) is a common problem with the mouth. People with gum disease might have bleeding gums around the teeth, swollen gums, receding gums, or sore or infected gums with symptoms lasting for at least 2 weeks and not caused by injury or problems with dentures. **Do you think you might have gum disease?**

□ Yes □ No

Have you ever had any adult teeth that became loose on their own, without some injury? (Milk teeth / baby teeth don’t count)

□ Yes □ No

Have you ever been told by a dental professional that you have lost bone around your teeth?

□ Yes □ No

How many teeth do you have – **excluding** wisdom teeth? (Usually adults have 28 non-wisdom teeth. Wisdom teeth are the four large teeth at the back of the jaw, that come through around the age of 20 – they are frequently extracted in the absence of other dental problems. Please **don’t** include them in your count.)

Display This Question:

If Q106_ud = Mental health disorder

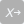

Q132 Have you ever been diagnosed with one or more of the following mental health disorders by a professional, even if you don’t have it currently? 

 
Please select all that apply:

- Depression (0)
- Depression during or after pregnancy (antenatal/postnatal depression) (1)
- Mania, hypomania, bipolar or manic-depression (2)
- Anxiety or generalised anxiety disorder (3)
- Social anxiety disorder or social phobia (4)
- Specific phobia (e.g. fear of flying) (5)
- Agoraphobia (6)
- Panic attacks or panic disorder (7)
- Post-traumatic stress disorder (PTSD) (8)
- Anorexia nervosa (9)
- Atypical anorexia nervosa (10)
- Bulimia nervosa (11)
- Binge-eating disorder (12)
- Other eating disorder (13)
- Obsessive-compulsive disorder (OCD) (14)
- Body Dysmorphic disorder (BDD) (15)
- Other obsessive-compulsive related disorders (e.g. skin-picking) (16)
- Schizophrenia (17)
- Any other type of psychotic illness (18)
- Personality disorder (19)
- Autism spectrum disorder (ASD) (20)
- Attention deficit or attention deficit and hyperactivity disorder (ADD/ADHD) (21)
- ⊗None of the above (22)

Skip To: Q136 If Condition: Have you ever been diagnose... Is Equal to 0. Skip To: Do you have any other long-standing h....

Skip To: Q136 If Q132 = None of the above

| Page Break |
| --- |

Display This Question:

If If Have you ever been diagnosed with one or more of the following mental health disorders by a professional, even if you don’t have it currently?&nbsp; &nbsp; Please select all that apply: q://QID179/SelectedChoicesCount Is Greater Than 0

And Q132 != None of the above

| 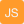 | 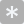 |
| --- | --- |

Q133 Please indicate below your **age at diagnosis** for the following mental health disorders you have been diagnosed with.

|  | Age at diagnosis |
| --- | --- |
|  | Age (1) |
| Depression (Q132_1) |  |
| Depression during or after pregnancy *(antenatal/postnatal depression)* (Q132_2) |  |
| Mania, hypomania, bipolar or manic-depression (Q132_3) |  |
| Anxiety or generalised anxiety disorder (Q132_4) |  |
| Social anxiety disorder or social phobia (Q132_5) |  |
| Specific phobia (e.g. fear of flying) (Q132_6) |  |
| Agoraphobia (Q132_7) |  |
| Panic attacks or panic disorder (Q132_8) |  |
| Post-traumatic stress disorder (PTSD) (Q132_9) |  |
| Anorexia nervosa (Q132_10) |  |
| Atypical anorexia nervosa (Q132_11) |  |
| Bulimia nervosa (Q132_12) |  |
| Binge-eating disorder (Q132_13) |  |
| Other eating disorder (Q132_14) |  |
| Obsessive-compulsive disorder (OCD) (Q132_15) |  |
| Body Dysmorphic disorder (BDD) (Q132_16) |  |
| Other obsessive-compulsive related disorders (e.g. skin-picking) (Q132_17) |  |
| Schizophrenia (Q132_18) |  |
| Any other type of psychotic illness (Q132_19) |  |
| Personality disorder (Q132_20) |  |
| Autism spectrum disorder (ASD) (Q132_21) |  |
| Attention deficit or attention deficit and hyperactivity disorder (ADD/ADHD) (Q132_22) |  |

| Page Break |
| --- |

Display This Question:

If Q132 != None of the above

And And Have you ever been diagnosed with one or more of the following mental health disorders by a professional, even if you don’t have it currently?&nbsp; &nbsp; Please select all that apply: q://QID179/SelectedChoicesCount Is Greater Than 0

| 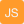 | 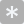 |
| --- | --- |

Q134
Please indicate which diagnoses you have received treatment for and if you are currently experiencing symptoms by selecting the box in the appropriate column.

|  | Have you received treatment? (e.g. seeking help or medication) | | Are you currently experiencing symptoms? | |
| --- | --- | --- | --- | --- |
|  | Yes (1) | No (2) | Yes (1) | No (2) |
| Depression (MHD.DX.1.1_1) |  |  |  |  |
| Depression during or after pregnancy *(antenatal/postnatal depression)* (MHD.DX.1.1_2) |  |  |  |  |
| Mania, hypomania, bipolar or manic-depression (Q182_17) |  |  |  |  |
| Anxiety or generalised anxiety disorder (Q182_18) |  |  |  |  |
| Social anxiety disorder or social phobia (Q182_19) |  |  |  |  |
| Specific phobia (e.g. fear of flying) (Q182_20) |  |  |  |  |
| Agoraphobia (Q182_21) |  |  |  |  |
| Panic attacks or panic disorder (Q182_22) |  |  |  |  |
| Post-traumatic stress disorder (PTSD) (Q182_23) |  |  |  |  |
| Anorexia nervosa (Q182_24) |  |  |  |  |
| Atypical anorexia nervosa (Q182_25) |  |  |  |  |
| Bulimia nervosa (Q182_26) |  |  |  |  |
| Binge-eating disorder (Q182_27) |  |  |  |  |
| Other eating disorder (Q182_28) |  |  |  |  |
| Obsessive-compulsive disorder (OCD) (Q182_29) |  |  |  |  |
| Body Dysmorphic disorder (BDD) (Q182_30) |  |  |  |  |
| Other obsessive-compulsive related disorders (e.g. skin-picking) (Q182_31) |  |  |  |  |
| Schizophrenia (Q182_32) |  |  |  |  |
| Any other type of psychotic illness (Q182_33) |  |  |  |  |
| Personality disorder (Q182_34) |  |  |  |  |
| Autism spectrum disorder (ASD) (Q182_35) |  |  |  |  |
| Attention deficit or attention deficit and hyperactivity disorder (ADD/ADHD) (Q182_36) |  |  |  |  |

| Page Break |
| --- |

Display This Question:

If Q132 = Personality disorder

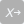

Q135 Which personality disorder have you been diagnosed with?

- Paranoid personality disorder (0)
- Narcissistic personality disorder (1)
- Schizoid personality disorder (2)
- Avoidant/anxious personality disorder (3)
- Dependent personality disorder (4)
- Schizotypal personality disorder (5)
- Antisocial personality disorder (6)
- Obsessive-compulsive personality disorder (7)
- Emotionally unstable personality disorder (Borderline personality disorder) (8)
- Histrionic personality disorder (9)
- Don’t know (10)
- Prefer not to answer (11)

| Page Break |
| --- |

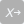

Q136 Do you have any other long-standing health condition or disability?

- Yes (1)
- No (0)

| Page Break |
| --- |

Display This Question:

If Q136 = Yes

Q137 If yes, please specify by entering the name of each condition, using one box per condition:

|  | Name of condition (1) |
| --- | --- |
| Condition 1 (4) |  |
| Condition 2 (5) |  |
| Condition 3 (6) |  |
| Condition 4 (7) |  |
| Condition 5 (8) |  |
| Condition 6 (9) |  |
| Condition 7 (10) |  |
| Condition 8 (11) |  |
| Condition 9 (12) |  |
| Condition 10 (13) |  |

| Page Break |
| --- |

Display This Question:

If Q136 = Yes

Q138 Please indicate below your **age at diagnosis** for each of the long-standing health conditions you listed previously:

|  | Age at Diagnosis (1) |
| --- | --- |
| If If yes, please specify by entering the name of each condition, using one box per condition: Condition 1 - Name of condition Is Not Empty  ${Q137/ChoiceTextEntryValue/4/1} (1) |  |
| If If yes, please specify by entering the name of each condition, using one box per condition: Condition 2 - Name of condition Is Not Empty  ${Q137/ChoiceTextEntryValue/5/1} (2) |  |
| If If yes, please specify by entering the name of each condition, using one box per condition: Condition 3 - Name of condition Is Not Empty  ${Q137/ChoiceTextEntryValue/6/1} (3) |  |
| If If yes, please specify by entering the name of each condition, using one box per condition: Condition 4 - Name of condition Is Not Empty  ${Q137/ChoiceTextEntryValue/7/1} (4) |  |
| If If yes, please specify by entering the name of each condition, using one box per condition: Condition 5 - Name of condition Is Not Empty  ${Q137/ChoiceTextEntryValue/8/1} (5) |  |
| If If yes, please specify by entering the name of each condition, using one box per condition: Condition 6 - Name of condition Is Not Empty  ${Q137/ChoiceTextEntryValue/9/1} (6) |  |
| If If yes, please specify by entering the name of each condition, using one box per condition: Condition 7 - Name of condition Is Not Empty  ${Q137/ChoiceTextEntryValue/10/1} (7) |  |
| If If yes, please specify by entering the name of each condition, using one box per condition: Condition 8 - Name of condition Is Not Empty  ${Q137/ChoiceTextEntryValue/11/1} (8) |  |
| If If yes, please specify by entering the name of each condition, using one box per condition: Condition 9 - Name of condition Is Not Empty  ${Q137/ChoiceTextEntryValue/12/1} (9) |  |
| If If yes, please specify by entering the name of each condition, using one box per condition: Condition 10 - Name of condition Is Not Empty  ${Q137/ChoiceTextEntryValue/13/1} (10) |  |

| Page Break |
| --- |

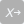

Q139 Have you had a **parent, a brother, a sister or a child** that has been diagnosed with a mental health problem? 

We are asking about **blood relatives only**, so information about relatives by adoption (e.g. adopted child, adoptive parents) should not be provided. We are **not** asking about more distant relatives (e.g. grandparents, cousins, grandchildren), or relatives by marriage (e.g. husband / wife).

- Yes (1)
- No (0)

| Page Break |
| --- |

Display This Question:

If Q139 = Yes

| 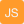 | 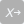 |
| --- | --- |

Q140 Please indicate which mental health problem your first-degree blood relatives (parents, siblings, children) have been diagnosed with. Select as many that may apply:

- Agoraphobia (0)
- Anorexia nervosa (1)
- Anxiety or generalised anxiety disorder (2)
- Attention deficit or attention deficit and hyperactivity disorder (ADD/ADHD) (3)
- Atypical anorexia nervosa (4)
- Autism spectrum disorder (ASD) (5)
- Binge-eating disorder (6)
- Body Dysmorphic disorder (BDD) (7)
- Bulimia nervosa (8)
- Depression (9)
- Depression during or after pregnancy *(antenatal/postnatal depression)* (10)
- Mania, hypomania, bipolar or manic-depression (11)
- Obsessive-compulsive disorder (OCD) (12)
- Panic attacks or panic disorder (13)
- Personality disorder (14)
- Post-traumatic stress disorder (PTSD) (15)
- Schizophrenia (16)
- Social anxiety disorder or social phobia (17)
- Specific phobia (e.g. fear of flying) (18)
- Any other type of psychotic illness (19)
- Other eating disorder (20)
- Other obsessive-compulsive related disorders (e.g. skin-picking) (21)
- ⊗Not sure (22)

End of Block: COVIDENCE_Long-standing Conditions

Start of Block: COVIDENCE_Medications and immunisations

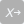

Q141 Do you currently take any prescribed medicines or treatments?

- Yes (1)
- No (0)

| Page Break |
| --- |

Display This Question:

If Q141 = Yes

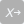

Q142 Please type the names of all the medications you are currently taking below, one medication per box. The next pages will collect details about dosage for each.

Include all types of medications taken at home or administered in a hospital or clinic (capsules, tablets, contraceptive pills or implants, inhalers, injections, intravenous infusions, monoclonal antibodies, chemotherapy, immunosuppressants, etc.)

Please note that other pages will collect details about the amount of medicine in each dose (next page) and how often you take each dose (the page after that). If there are any details about your medication that aren’t captured by our form (e.g. if you take different doses of a medicine at different times of day), there will be space to enter them in a blank text box at the end of this section of the questionnaire.

|  | Medication name |
| --- | --- |
|  | (Generic or Brand name, either is fine) (1) |
| Medication 1 (2) |  |
| Medication 2 (3) |  |
| Medication 3 (4) |  |
| Medication 4 (5) |  |
| Medication 5 (6) |  |
| Medication 6 (7) |  |
| Medication 7 (8) |  |
| Medication 8 (9) |  |
| Medication 9 (10) |  |
| Medication 10 (11) |  |
| Medication 11 (12) |  |
| Medication 12 (13) |  |
| Medication 13 (14) |  |
| Medication 14 (15) |  |
| Medication 15 (16) |  |
| Medication 16 (17) |  |
| Medication 17 (18) |  |
| Medication 18 (19) |  |
| Medication 19 (20) |  |
| Medication 20 (21) |  |

| Page Break |
| --- |

Display This Question:

If Q141 = Yes

Q143 Please enter the amount of medicine that is in each dose you take in the table below. Note that information about how often you take that dose will be asked in the next question. For example, if you take a 20 mg tablet twice per day, enter 20 mg in this table – and twice per day in the table after this one.

If you do not know the dose, please enter “0” in the box.

|  | Dose | Dose units |
| --- | --- | --- |
|  | (1) |  |
| ${Q142%231/ChoiceTextEntryValue/2/1} (2) |  | ▼ milligrams (mg) (1 ... other (6) |
| ${Q142%231/ChoiceTextEntryValue/3/1} (3) |  | ▼ milligrams (mg) (1 ... other (6) |
| ${Q142%231/ChoiceTextEntryValue/4/1} (4) |  | ▼ milligrams (mg) (1 ... other (6) |
| ${Q142%231/ChoiceTextEntryValue/5/1} (5) |  | ▼ milligrams (mg) (1 ... other (6) |
| ${Q142%231/ChoiceTextEntryValue/6/1} (6) |  | ▼ milligrams (mg) (1 ... other (6) |
| ${Q142%231/ChoiceTextEntryValue/7/1} (7) |  | ▼ milligrams (mg) (1 ... other (6) |
| ${Q142%231/ChoiceTextEntryValue/8/1} (8) |  | ▼ milligrams (mg) (1 ... other (6) |
| ${Q142%231/ChoiceTextEntryValue/9/1} (9) |  | ▼ milligrams (mg) (1 ... other (6) |
| ${Q142%231/ChoiceTextEntryValue/10/1} (10) |  | ▼ milligrams (mg) (1 ... other (6) |
| ${Q142%231/ChoiceTextEntryValue/11/1} (11) |  | ▼ milligrams (mg) (1 ... other (6) |
| ${Q142%231/ChoiceTextEntryValue/12/1} (12) |  | ▼ milligrams (mg) (1 ... other (6) |
| ${Q142%231/ChoiceTextEntryValue/13/1} (13) |  | ▼ milligrams (mg) (1 ... other (6) |
| ${Q142%231/ChoiceTextEntryValue/14/1} (14) |  | ▼ milligrams (mg) (1 ... other (6) |
| ${Q142%231/ChoiceTextEntryValue/15/1} (15) |  | ▼ milligrams (mg) (1 ... other (6) |
| ${Q142%231/ChoiceTextEntryValue/16/1} (16) |  | ▼ milligrams (mg) (1 ... other (6) |
| ${Q142%231/ChoiceTextEntryValue/17/1} (17) |  | ▼ milligrams (mg) (1 ... other (6) |
| ${Q142%231/ChoiceTextEntryValue/18/1} (18) |  | ▼ milligrams (mg) (1 ... other (6) |
| ${Q142%231/ChoiceTextEntryValue/19/1} (19) |  | ▼ milligrams (mg) (1 ... other (6) |
| ${Q142%231/ChoiceTextEntryValue/20/1} (20) |  | ▼ milligrams (mg) (1 ... other (6) |
| ${Q142%231/ChoiceTextEntryValue/21/1} (21) |  | ▼ milligrams (mg) (1 ... other (6) |

| Page Break |
| --- |

Display This Question:

If Q141 = Yes

Q144 Please select the frequency and route that you take your medication:

|  | Frequency | Route |
| --- | --- | --- |
| ${Q142%231/ChoiceTextEntryValue/2/1} (2) | ▼ 4 times/day (1 ... As needed (7) | ▼ by mouth (1 ... other (4) |
| ${Q142%231/ChoiceTextEntryValue/3/1} (3) | ▼ 4 times/day (1 ... As needed (7) | ▼ by mouth (1 ... other (4) |
| ${Q142%231/ChoiceTextEntryValue/4/1} (4) | ▼ 4 times/day (1 ... As needed (7) | ▼ by mouth (1 ... other (4) |
| ${Q142%231/ChoiceTextEntryValue/5/1} (5) | ▼ 4 times/day (1 ... As needed (7) | ▼ by mouth (1 ... other (4) |
| ${Q142%231/ChoiceTextEntryValue/6/1} (6) | ▼ 4 times/day (1 ... As needed (7) | ▼ by mouth (1 ... other (4) |
| ${Q142%231/ChoiceTextEntryValue/7/1} (7) | ▼ 4 times/day (1 ... As needed (7) | ▼ by mouth (1 ... other (4) |
| ${Q142%231/ChoiceTextEntryValue/8/1} (8) | ▼ 4 times/day (1 ... As needed (7) | ▼ by mouth (1 ... other (4) |
| ${Q142%231/ChoiceTextEntryValue/9/1} (9) | ▼ 4 times/day (1 ... As needed (7) | ▼ by mouth (1 ... other (4) |
| ${Q142%231/ChoiceTextEntryValue/10/1} (10) | ▼ 4 times/day (1 ... As needed (7) | ▼ by mouth (1 ... other (4) |
| ${Q142%231/ChoiceTextEntryValue/11/1} (11) | ▼ 4 times/day (1 ... As needed (7) | ▼ by mouth (1 ... other (4) |
| ${Q142%231/ChoiceTextEntryValue/12/1} (13) | ▼ 4 times/day (1 ... As needed (7) | ▼ by mouth (1 ... other (4) |
| ${Q142%231/ChoiceTextEntryValue/13/1} (14) | ▼ 4 times/day (1 ... As needed (7) | ▼ by mouth (1 ... other (4) |
| ${Q142%231/ChoiceTextEntryValue/14/1} (15) | ▼ 4 times/day (1 ... As needed (7) | ▼ by mouth (1 ... other (4) |
| ${Q142%231/ChoiceTextEntryValue/15/1} (16) | ▼ 4 times/day (1 ... As needed (7) | ▼ by mouth (1 ... other (4) |
| ${Q142%231/ChoiceTextEntryValue/16/1} (17) | ▼ 4 times/day (1 ... As needed (7) | ▼ by mouth (1 ... other (4) |
| ${Q142%231/ChoiceTextEntryValue/17/1} (18) | ▼ 4 times/day (1 ... As needed (7) | ▼ by mouth (1 ... other (4) |
| ${Q142%231/ChoiceTextEntryValue/18/1} (19) | ▼ 4 times/day (1 ... As needed (7) | ▼ by mouth (1 ... other (4) |
| ${Q142%231/ChoiceTextEntryValue/19/1} (20) | ▼ 4 times/day (1 ... As needed (7) | ▼ by mouth (1 ... other (4) |
| ${Q142%231/ChoiceTextEntryValue/20/1} (21) | ▼ 4 times/day (1 ... As needed (7) | ▼ by mouth (1 ... other (4) |
| ${Q142%231/ChoiceTextEntryValue/21/1} (22) | ▼ 4 times/day (1 ... As needed (7) | ▼ by mouth (1 ... other (4) |

| Page Break |
| --- |

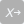

Q145 Do you regularly take tablets containing any of the following painkillers or anti-inflammatories?  (e.g. for joint pain, back pain, period pain etc.) 

Select as many as apply.

- Paracetamol (0)
- Ibuprofen (1)
- Diclofenac (2)
- Naproxen (3)
- Aspirin (4)
- ⊗None of the above (5)

| Page Break |
| --- |

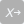

Q146 Are there any medicines (e.g. immunosuppressants) that you used to take regularly before the coronavirus outbreak, but which have been stopped by your doctor in order to reduce your risk of COVID-19?

- Yes (1)
- No (0)

| Page Break |
| --- |

Display This Question:

If Q146 = Yes

Q147 If yes, please list them here:

________________________________________________________________

________________________________________________________________

________________________________________________________________

________________________________________________________________

________________________________________________________________

| Page Break |
| --- |

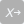

Q148 Is there is any detail of your medication that we have not been able to capture in the tables above (e.g. if you take different doses of a particular medicine at different times of day)?

- Yes (1)
- No (0)

| Page Break |
| --- |

Display This Question:

If Q148 = Yes

Q149 If yes, please provide details here:

________________________________________________________________

________________________________________________________________

________________________________________________________________

________________________________________________________________

________________________________________________________________

| Page Break |
| --- |

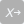

Q149.1 Have you had one or more doses of a COVID-19 vaccine (immunisation)? First doses and booster doses both count.

- Yes (1)
- No (2)
- Not sure (e.g. you took part in vaccine trial, but don’t yet know whether or not you had the real vaccine or the placebo (dummy vaccine) (3)

| Page Break |
| --- |

Display This Question:

If Q149.1= Yes

Q149.2 How many doses of COVID-19 vaccine have you had in total?

- One dose (1)
- Two doses (2)

| Page Break |
| --- |

Display This Question:

If Q149.2 = One dose

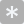

Q149.3 On what date did you have this vaccine dose? If you are not sure of the exact date, enter the approximate date (DD/MM/YYYY) e.g. for 16th December 2020, write 16/12/2020

________________________________________________________________

| Page Break |
| --- |

Display This Question:

If Q149.2= Two doses

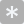

Q149.4 On what date did you have the first of these vaccine doses? If you are not sure of the exact date, enter the approximate date (DD/MM/YYYY) e.g. for 16th December 2020, write 16/12/2020

________________________________________________________________

Display This Question:

If Q149.2 = Two doses

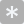

Q149.5 On what date did you have the second of these vaccine doses? If you are not sure of the exact date, enter the approximate date (DD/MM/YYYY) e.g. for 16th December 2020, write 16/12/2020

________________________________________________________________

| Page Break |
| --- |

Display This Question:

If Q149.1 = Yes

Q149.6 Which COVID-19 vaccine did you have?

- Oxford / AstraZeneca / ChAdOx1 (1)
- Pfizer / BioNTech (2)
- Moderna (3)
- Novavax (4)
- Other - please specify (5) ________________________________________________
- Not sure / don't know (6)

Display This Question:

If Q149.1 = Yes

Q149.7

Did you experience any of the following symptoms following your COVID-19 vaccine?

- Tenderness, soreness, swelling, redness or a painful, heavy feeling at the injection site (0)
- Feeling tired (1)
- Headache (2)
- Fever / high temperature (37.8C or greater) (3)
- Muscle aches (4)
- swelling of the glands in your armpit or neck (5)
- other symptoms: specify (6)
- none of the above (7)

Display This Question:

If Q149.1 = Yes

Q149.8

Did you take any of the following to treat post-vaccination symptoms (e.g. fever, muscle aches etc) ?

- Paracetamol (e.g. Disprol, Hedex, Medinol, Panadol) (1)
- Ibuprofen (e.g. Nurofen, Brufen, Calprofen) (2)
- Something else: please write name here (3)
- None of the above (4)

Q150 Have you had a flu vaccine in the previous 12 months?

- Yes (1)
- No (0)

| Page Break |
| --- |

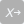

Q151 Have you had a pneumococcal vaccine in the previous 12 months?

- Yes (1)
- No (0)

| Page Break |
| --- |

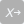

Q152 Have you ever had the BCG vaccine?

*This is the vaccine against Tuberculosis (TB), it's injected in the upper arm and usually leaves a small scar.*

- Yes (1)
- No (0)
- Unsure (2)

| Page Break |
| --- |

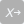

Q153 Have you ever had the MMR vaccine?

*This is the vaccine against measles, mumps and rubella.*

- Yes (1)
- No (0)
- Unsure (2)

End of Block: COVIDENCE_Medications and immunisations

Start of Block: COVIDENCE_Mental well-being

Q154 We would now like to ask a few questions about your mental well-being.

Our mental health, particularly depression and anxiety, is closely entwined with our physical health and can play a role in how well we fight an illness or respond to treatment. That’s why we want to ask these questions.

In the last week, how often have you been bothered by the following problems?

|  | Not at all (1) | Several days (1-3 days/week) (2) | More than half the days (4-5 days/week) (3) | Nearly every day (6-7 days/week) (4) |
| --- | --- | --- | --- | --- |
| Little interest or pleasure in doing things (1) |  |  |  |  |
| Feeling down, depressed, or hopeless (2) |  |  |  |  |
| Feeling nervous, anxious or on edge (3) |  |  |  |  |
| Not being able to stop or control worrying (5) |  |  |  |  |

| Page Break |
| --- |

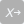

Q155 **Over the past month,** have you noticed any new and unusual experiences – like hearing things or seeing things that others could not or when other people are not present?

- Yes (1)
- No (0)

| Page Break |
| --- |

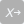

Q156 In the past 2 weeks, how often have you been bothered by unpleasant thoughts, urges, or images that repeatedly enter your mind?

- Nearly every day (0)
- More than half the days (1)
- Several days (2)
- Rare, less than a day or two (3)
- Not at all (4)

| Page Break |
| --- |

End of Block: COVIDENCE_Mental well-being

Start of Block: COVIDENCE_Senses

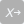

Q157
We now would like to ask a few questions about your sense of smell, your sense of taste and your hearing.

Please make an active attempt to smell any one of the following: orange, coffee, vinegar, Vicks, Dettol.

Over the last month, have you noticed any loss in your sense of smell or your ability to taste food?

- Yes (1)
- No (0)

| Page Break |
| --- |

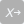

Q158 Over the last month, have you had noises (such as ringing or buzzing) in your head or in one or both ears that lasted for more than five minutes at a time?

- Yes (1)
- No (0)

End of Block: COVIDENCE_Senses

Start of Block: COVIDENCE_Response to Coronavirus Outbreak

Q159 Finally, we would like to ask some questions about your lifestyle that might affect risk of coronavirus infection.

| Page Break |
| --- |

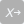

Q160 In the last week, and on average, how many times per day did you wash your hands with soap and water or use hand sanitiser gel?

▼ 0 (0) ... 10 or more (10)

| Page Break |
| --- |

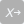

Q161 In the last week, how many journeys did you make on public transport? (return trips count as 2 journeys)

▼ 0 (0) ... 10 or more (10)

| Page Break |
| --- |

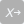

Q162 In the last week, how frequently have you worn a face mask while in an indoor public place?

- Always (100% of the time) (0)
- Usually (50%-99%) (1)
- Sometimes (1-49% of the time) (2)
- Never (0% of the time) (3)
- Not applicable (I haven't been in a public place in the last week) (4)

| Page Break |
| --- |

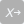

Q163 In the last week, how frequently have you worn gloves as an infection protection measure while outside the home?

- Always (100% of the time) (0)
- Usually (50%-99%) (1)
- Sometimes (1-49% of the time) (2)
- Never (0% of the time) (3)
- Not applicable (I haven't been in a public place in the last week) (4)

| Page Break |
| --- |

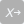

Q164 Over the last week, how many days did you work/study exclusively from home?

▼ 0 (0) ... I am not currently working or studying (9)

| Page Break |
| --- |

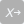

Q165 Over the last week, how many times have you been visited at home by someone who does not live in your household? 

(We are referring to visitors entering inside your home. People who just call at the door and do not come inside do not count)

▼ 0 (0) ... 10 or more (10)

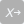

Q166 Over the last week, how many times have you been into the home of someone who does not live in your household? 

(We are referring to visits in which you enter inside someone else’s home. Visits where you do not cross the threshold do not count. Neither do visits to the shops or other public places, we will ask about these later.)

▼ 0 (0) ... 10 or more (10)

| Page Break |
| --- |

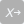

Q167 In the last week, how many times have you been inside a shop or supermarket?

▼ 0 (0) ... 10 or more (10)

| Page Break |
| --- |

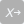

Q168 In the last week, how often have you been inside another indoor public space (e.g. café, pub, place of worship, restaurant, gym, day centre, waiting room, school, library, entertainment venue, hairdresser, takeaway restaurant)?

▼ 0 (0) ... 10 or more (10)

| Page Break |
| --- |

Q168.1 In the last month, have any ‘local restrictions’ (i.e. over and above national restrictions) been put in place in the area where you live as a result of a local increase in COVID-19 cases? Examples include restrictions on social contact and/or travel, and temporary closure of local businesses or venues.

- Yes (1)
- No (2)
- Not sure (3)

| Page Break |
| --- |

Q168.2 Have you travelled outside of the UK since February 2020?

- Yes (1)
- No (2)

| Page Break |
| --- |

Display This Question:

If Q168.2 = Yes

Q168.3 If yes, please write the name of the country or countries you have visited here:

________________________________________________________________

| Page Break |
| --- |

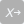

Q169 Is there any other information you think would be useful to us, or any clarification you would like to make regarding the information you have provided today?

- Yes (1)
- No (0)

| Page Break |
| --- |

Display This Question:

If Q169 = Yes

Q170 If yes, please provide details:

________________________________________________________________

________________________________________________________________

________________________________________________________________

________________________________________________________________

________________________________________________________________

Q171 Finally, can you let us know how you first heard about COVIDENCE UK?  This question is optional, but your answer will help us work out the most effective way to spread the word about the study.
Click as many as apply (e.g. if you heard about the study from a friend via twitter, click both boxes)

- TV (1)
- Radio (2)
- Newspaper (paper copy or newspaper website) (3)
- Facebook (4)
- Instagram (5)
- Twitter (6)
- Linkedin (7)
- From a friend or family member (8)
- Poster (9)
- Flyer/leaflet (10)
- Printed advertisement in magazine (11)
- Online advertisement (12)
- From a charity (e.g. Diabetes UK, British Heart Foundation, British Lung Foundation etc), via newsletter or email (13)
- Via email from Discover (NW London Health Research Registrar) (14)

End of Block: COVIDENCE_Response to Coronavirus Outbreak

### COVIDENCE UK Monthly Questionnaire

Welcome to your monthly questionnaire for the COVIDENCE UK study.
  
Please select your answers carefully. Questions that only require one answer will automatically forward you to the next question. If you feel that there is important additional information that has not been captured by our questionnaire, you can enter this into the free text box provided at the end of the survey.
  
First, we will ask about the results of any recent nose/throat swab tests you have had since you last checked in with us.

| Page Break |
| --- |

End of Block: COVIDENCE_Intro

Start of Block: COVIDENCE_Recent Health

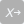

Q1 **Since you last checked in with us (last questionnaire sent [add date])**, have you had one or more nose or throat swabs for COVID-19 or any other respiratory virus, or have results from a previous swab test become newly available? (This question is about tests to detect the virus itself: they are usually done in somebody who has symptoms, but screening of asymptomatic people can also be done.)

- Yes (1)
- No (0)

| Page Break |
| --- |

Display This Question:

If Since you last checked in with us (last questionnaire sent [add date]), have you had one or mo... = Yes

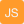

Q2_ud_v3 On what date did you have this nose / throat swab? Please select a date on the calendar below. 

 Only one date should be selected. If you are not sure of the exact date, click on the approximate date.

 If you had one or more positive swabs, **please select the date of your first positive swab**.

 If you have had multiple negative swabs, **please select the date of your first negative swab**.

________________________________________________________________

| Page Break |
| --- |

Display This Question:

If Since you last checked in with us (last questionnaire sent [add date]), have you had one or mo... = Yes

Q3_ud What was the result? Click as many as apply.

- Positive for COVID-19 (SARS-CoV-2 coronavirus) (1)
- Positive for influenza virus (2)
- Positive for another respiratory virus (3)
- Negative for all/any viruses tested (4)
- Not Known (5)

| Page Break |
| --- |

M2.15_ud **Since you last checked in with us (last questionnaire sent [add date])**, have you had one or more doses of a COVID-19 vaccine (immunisation)? First doses and booster doses both count.

 (You should only click ‘Yes’ if you have had a vaccine dose since you last completed a study questionnaire. If there are earlier vaccine doses that you have previously told us about, there is no need to enter their details again.)

- Yes, I have had one or more doses of COVID-19 vaccine **since I filled my last questionnaire** (1)
- No, I have not had any doses of COVID-19 vaccine **since I filled my last questionnaire**  (2)
- Not sure (e.g. you took part in vaccine trial, but don’t yet know whether or not you had the real vaccine or the placebo (dummy vaccine) (3)

| Page Break |
| --- |

Display This Question:

If Since you last checked in with us (last questionnaire sent [add date]), have you had one or mo... = Yes, I have had one or more doses of COVID-19 vaccine <strong>since I filled my last questionnaire</strong>

M2.16.1 How many doses of COVID-19 vaccine have you had since you last checked in with us? (last questionnaire sent [add date])

- One dose **since my last questionnaire** (1)
- Two doses **since my last questionnaire** (2)

| Page Break |
| --- |

Display This Question:

If How many doses of COVID-19 vaccine have you had since you last checked in with us? (last question... = One dose <strong>since my last questionnaire</strong>

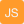

M2.16.2_ud_v3 On what date did you have this vaccine dose? Please select a date on the calendar below. 

 If you are not sure of the exact date, click on the approximate date.

________________________________________________________________

| Page Break |
| --- |

Display This Question:

If How many doses of COVID-19 vaccine have you had since you last checked in with us? (last question... = Two doses <strong>since my last questionnaire</strong>

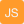

Q374_ud_v3 On what date did you have the first of these vaccine doses? Please select a date on the calendar below. 

 If you are not sure of the exact date, click on the approximate date.

________________________________________________________________

| Page Break |
| --- |

Display This Question:

If How many doses of COVID-19 vaccine have you had since you last checked in with us? (last question... = Two doses <strong>since my last questionnaire</strong>

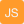

Q375_ud_v3 On what date did you have the second of these vaccine doses?  Please select a date on the calendar below. 

 If you are not sure of the exact date, click on the approximate date.

________________________________________________________________

| Page Break |
| --- |

Display This Question:

If Since you last checked in with us (last questionnaire sent [add date]), have you had one or mo... = Yes, I have had one or more doses of COVID-19 vaccine <strong>since I filled my last questionnaire</strong>

M2.16.3 Which COVID-19 vaccine did you have?

- Oxford / AstraZeneca / ChAdOx1 (1)
- Pfizer / BioNTech (2)
- Moderna (3)
- Valneva (7)
- Novavax (4)
- Janssen (also known as Johnson & Johnson) (8)
- Other - please specify (5) ________________________________________________
- Not sure / don't know (6)

| Page Break |
| --- |

Display This Question:

If Since you last checked in with us (last questionnaire sent [add date]), have you had one or mo... = Yes, I have had one or more doses of COVID-19 vaccine <strong>since I filled my last questionnaire</strong>

Or Since you last checked in with us (last questionnaire sent [add date]), have you had one or mo... = Not sure (e.g. you took part in vaccine trial, but don’t yet know whether or not you had the real vaccine or the placebo (dummy vaccine)

M2.16.31 Did you experience any of the following symptoms following your COVID-19 vaccine?

- Tenderness, soreness, swelling, redness or a painful, heavy feeling at the injection site (1)
- Feeling tired (2)
- Headache (3)
- Fever / high temperature (37.8° C or greater) (4)
- Muscle aches (5)
- Swelling of the glands in your armpit or neck (6)
- Other symptoms: specify (7) ________________________________________________
- ⊗None of the above (8)

| Page Break |
| --- |

Display This Question:

If Since you last checked in with us (last questionnaire sent [add date]), have you had one or mo... = Yes, I have had one or more doses of COVID-19 vaccine <strong>since I filled my last questionnaire</strong>

Or Since you last checked in with us (last questionnaire sent [add date]), have you had one or mo... = Not sure (e.g. you took part in vaccine trial, but don’t yet know whether or not you had the real vaccine or the placebo (dummy vaccine)

M2.16.32 Did you take any of the following to treat post-vaccination symptoms (e.g. fever, muscle aches etc)?

- Paracetamol (e.g. Disprol, Hedex, Medinol, Panadol) (1)
- Ibuprofen (e.g. Nurofen, Brufen, Calprofen) (2)
- Something else: please write name here (3) ________________________________________________
- ⊗None of the above (5)

| Page Break |
| --- |

M2.17 **Since you last checked in with us (last questionnaire sent [add date]),** have you had a flu jab (vaccine against influenza)?

- Yes (1)
- No (2)

| Page Break |
| --- |

Display This Question:

If Since you last checked in with us (last questionnaire sent [add date]), have you had a flu jab... = Yes

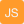

M1.18_ud_v3 On what date did you have this flu jab (immunisation)? Please select a date on the calendar below. 

 If you are not sure of the exact date, click on the approximate date.

________________________________________________________________

| Page Break |
| --- |

M0 *IMPORTANT NOTE: The following question is designed to capture symptoms of ANY and ALL respiratory infections, i.e. common cold, chest infections and ‘flu - as well as COVID-19 of course.

 Please click ‘yes’ if you have had ANY symptoms of respiratory infection – cold symptoms and sore throat count as well as classic COVID-19 symptoms (loss of smell/taste, fever and persistent cough). Your answers to the detailed questions about the nature of your symptoms will allow us to tell the different infections apart.* 
 
 

M3_ud **Since you last checked in with us (last questionnaire sent [add date])**, have you experienced any of the following symptoms: cold or flu symptoms, sore throat, persistent cough, loss of smell or taste, fever, fatigue, diarrhoea, abdominal pain or loss of appetite?

- Yes, I have had one or more of these symptoms since completing my last COVIDENCE UK questionnaire (1)
- No, I have not had any of these symptoms since completing my last COVIDENCE UK questionnaire (2)

Skip To: M3.2 If Since you last checked in with us (last questionnaire sent [add date]), have you experienced a... = No, I have not had any of these symptoms since completing my last COVIDENCE UK questionnaire

| Page Break |
| --- |

Display This Question:

If Since you last checked in with us (last questionnaire sent [add date]), have you experienced a... = Yes, I have had one or more of these symptoms since completing my last COVIDENCE UK questionnaire

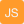

M3.1.1_ud_v3 When did your symptoms start? Please select a date on the calendar below. 

 If you are not sure of the exact date, click on the approximate date. 

________________________________________________________________

| Page Break |
| --- |

M3.1.2 How ill did you feel at your worst?

- Mildly unwell - I could do most of my usual activities (1)
- Moderately unwell - I couldn’t do usual activities, but didn’t need to go to bed in the daytime (2)
- Very unwell – I had to go to bed in the daytime (3)

| Page Break |
| --- |

M3.1.3 Did you have a fever (high temperature)?

- Yes (1)
- No (2)

| Page Break |
| --- |

Display This Question:

If Did you have a fever (high temperature)? = Yes

M3.1.3.1 What was your highest temperature?

- I don’t know / I did not take my temperature (1)
- Less than 38°C (less than 100°F) (2)
- 38.0-39.0°C (100-102°F) (3)
- More than 39°C (more than 102°F) (4)

| Page Break |
| --- |

M3.1.4 Did you have a persistent cough (coughing a lot for more than an hour, or 3 or more coughing episodes in 24 hours)?

- No (1)
- Persistent dry cough (i.e. producing little or no phlegm) (2)
- Persistent productive cough (3)

| Page Break |
| --- |

M3.1.5 Did you experience unusual fatigue?

- No (1)
- Mild fatigue (2)
- Severe fatigue - I struggled to get out of bed (3)

| Page Break |
| --- |

M3.1.6 Did you have a headache?

- Yes (1)
- No (2)

| Page Break |
| --- |

M3.1.7 Did you have muscle aches?

- Yes (1)
- No (2)

| Page Break |
| --- |

M3.1.7.5 Did you have a loss of sense of smell or taste?

- Yes (1)
- No (2)

| Page Break |
| --- |

M3.1.8  Did you experience unusual shortness of breath?

- No (1)
- Yes, mild symptoms - slight shortness of breath during ordinary activity (2)
- Yes, significant symptoms – breathing was comfortable only at rest (3)
- Yes, severe symptoms - breathing was difficult even at rest (4)

| Page Break |
| --- |

| 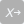 | 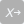 |
| --- | --- |

M3.1.9 Please indicate any other symptoms you had. 

Please select as many as apply:

- Sore throat (1)
- Sneezing (2)
- Runny nose (3)
- Blocked nose (4)
- Unusually hoarse voice (5)
- Unusual chest pain (6)
- Unusual abdominal pain (7)
- Diarrhoea (8)
- Confusion, disorientation, or drowsiness (9)
- Raised, red itchy welts on the skin or sudden swelling of the face or lips (12)
- Red/purple sores or blisters on your feet or toes (13)
- Unusual soreness or discomfort of the eyes (e.g. light sensitivity, excessive tears, pink/red eyes) (14)
- Other symptom (please specify) (11) ________________________________________________
- ⊗None of the above (10)

| Page Break |
| --- |

M3.1.10 Did you skip any meals because you felt unwell?

- Yes (1)
- No (2)

| Page Break |
| --- |

M3.1.20 Did you take any over the counter medication to relieve your symptoms?

- Yes (1)
- No (2)

| Page Break |
| --- |

Display This Question:

If Did you take any over the counter medication to relieve your symptoms? = Yes

M3.1.20.1 You stated that you took over-the-counter medication, please click as many as apply:

- Paracetamol tablets (1)
- Cold and flu relief tablets or medicine (2)
- Ibuprofen (3)
- Naproxen (4)
- Aspirin (5)
- Other (6)

| Page Break |
| --- |

M3.1.205 Did you have a telephone appointment with your GP to discuss these symptoms?

- Yes (1)
- No (2)

| Page Break |
| --- |

M3.1.206 Did you have a face-to-face appointment with your GP to discuss these symptoms?

- Yes (1)
- No (2)

| Page Break |
| --- |

M3.1.207 Were you prescribed a course of antibiotics to treat these symptoms?

- Yes (1)
- No (2)

| Page Break |
| --- |

Display This Question:

If Did you have a telephone appointment with your GP to discuss these symptoms? = Yes

Or Did you have a face-to-face appointment with your GP to discuss these symptoms? = Yes

M3.1.208 What did the GP diagnose? Tick as many as apply

- Suspected or proven COVID-19 (1)
- Pneumonia (2)
- ‘Flu' (influenza) (3)
- Bronchitis (4)
- Tonsillitis or pharyngitis (sore throat) (5)
- Ear infection (otitis media) (6)
- Common cold (7)
- Another upper respiratory infection (8)
- Another lower respiratory infection (9)
- Something else (10)

| Page Break |
| --- |

Display This Question:

If What did the GP diagnose? Tick as many as apply = Something else

M3.1.209 Please write the diagnosis here if "something else":

________________________________________________________________

| Page Break |
| --- |

M3.1.21_ud Did you go to hospital for treatment of these symptoms?

- Yes, and I was admitted to hospital (i.e. I spent one or more nights as a hospital in-patient) (1)
- Yes, I attended a hospital accident and emergency department but I was not admitted to hospital (i.e. I went home without spending one or more nights as a hospital in-patient) (2)
- No, I didn’t go to hospital for treatment of these symptoms (3)

Skip To: M3.1.22 If Did you go to hospital for treatment of these symptoms? = No, I didn’t go to hospital for treatment of these symptoms

| Page Break |
| --- |

Display This Question:

If Did you go to hospital for treatment of these symptoms? = Yes, and I was admitted to hospital (i.e. I spent one or more nights as a hospital in-patient)

Or Did you go to hospital for treatment of these symptoms? = Yes, I attended a hospital accident and emergency department but I was not admitted to hospital (i.e. I went home without spending one or more nights as a hospital in-patient)

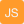

M3.1.21.1_ud_v3 On what date did you attend hospital? Please select a date on the calendar below. 

 If you are not sure of the exact date, click on the approximate date.

________________________________________________________________

| Page Break |
| --- |

M3.1.21.2 What treatment did you receive while in the hospital? 
Please select as many that apply:

- Oxygen and fluids (breathing support administered through an oxygen mask, no pressure applied) (2)
- Non-invasive ventilation (breathing support administered through an oxygen mask, which pushes oxygen into your lungs) (3)
- Invasive ventilation (breathing support administered through an inserted tube, people are usually asleep during this procedure) (4)
- I'm not sure (6)
- ⊗None (5)

| Page Break |
| --- |

M3.1.21.3 Have you been discharged from hospital?

- Yes (1)
- No (2)

| Page Break |
| --- |

Display This Question:

If Have you been discharged from hospital? = Yes

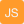

M3.1.21.3.1_ud_v3 On what date were you discharged from hospital? Please select a date on the calendar below. 
 
If you are not sure of the exact date, click on the approximate date.

________________________________________________________________

| Page Break |
| --- |

Q470 What did the hospital doctors diagnose? Select as many as apply.

- Suspected or proven COVID-19 (1)
- Pneumonia (2)
- ‘Flu' (influenza) (3)
- Bronchitis (4)
- Tonsillitis or pharyngitis (sore throat) (5)
- Ear infection (otitis media) (6)
- Common cold (7)
- Another upper respiratory infection (8)
- Another lower respiratory infection (9)
- Asthma attack (flare-up or exacerbation of asthma symptoms) (10)
- COPD attack (flare-up or exacerbation of COPD symptoms) (11)
- Something else (12)

M3.1.21.4_ud What did the hospital doctors diagnose? Select as many as apply.

- Suspected or proven COVID-19 (1)
- Pneumonia (2)
- ‘Flu' (influenza) (3)
- Bronchitis (4)
- Tonsillitis or pharyngitis (sore throat) (5)
- Ear infection (otitis media) (6)
- Common cold (7)
- Another upper respiratory infection (8)
- Another lower respiratory infection (9)
- Asthma attack (flare-up or exacerbation of asthma symptoms) (10)
- COPD attack (flare-up or exacerbation of COPD symptoms) (11)
- Something else (12)

Q471 What did the hospital doctors diagnose? Select as many as apply.

- Suspected or proven COVID-19 (1)
- Pneumonia (2)
- ‘Flu' (influenza) (3)
- Bronchitis (4)
- Tonsillitis or pharyngitis (sore throat) (5)
- Ear infection (otitis media) (6)
- Common cold (7)
- Another upper respiratory infection (8)
- Another lower respiratory infection (9)
- Asthma attack (flare-up or exacerbation of asthma symptoms) (10)
- COPD attack (flare-up or exacerbation of COPD symptoms) (11)
- Something else (12)

| Page Break |
| --- |

| Page Break |
| --- |

Display This Question:

If What did the hospital doctors diagnose? Select as many as apply.  = Something else

M3.1.21.4.1 If "something else", please write the diagnosis here:

________________________________________________________________

| Page Break |
| --- |

M3.1.22 Do you feel back to normal now?

- Yes (1)
- No (2)

| Page Break |
| --- |

Display This Question:

If Do you feel back to normal now? = Yes

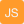

M3.1.22.1_ud_v3 On what date did you first feel back to normal? Please select a date on the calendar below. 

 If you are not sure of the exact date, click on the approximate date.

________________________________________________________________

| Page Break |
| --- |

M3.2 **Since you last checked in with us**, have you had an attack (exacerbation) of asthma or COPD (chronic bronchitis / emphysema)?

- Yes (1)
- No (4)

| Page Break |
| --- |

Display This Question:

If Since you last checked in with us, have you had an attack (exacerbation) of asthma or COPD (chron... = Yes

M3.21 Was this an attack of asthma or COPD?

- Asthma attack (1)
- COPD attack (4)

| Page Break |
| --- |

Display This Question:

If Since you last checked in with us, have you had an attack (exacerbation) of asthma or COPD (chron... = Yes

M3.22 Did this asthma/COPD attack require treatment with steroid tablets (e.g. prednisolone)?

- Yes (1)
- No (2)
- Don't know / not sure (3)

| Page Break |
| --- |

Display This Question:

If Since you last checked in with us, have you had an attack (exacerbation) of asthma or COPD (chron... = Yes

M3.23 Did this asthma/COPD attack require treatment with antibiotics (e.g. amoxycillin, clarithromycin, augmentin)?

- Yes (1)
- No (2)
- Don't know / not sure (3)

| Page Break |
| --- |

Display This Question:

If Since you last checked in with us, have you had an attack (exacerbation) of asthma or COPD (chron... = Yes

M3.24 Did you go to hospital for treatment of your asthma/COPD attack?

- No, I didn’t go to hospital for treatment of my attack (1)
- Yes, I attended a hospital accident and emergency department but I was not admitted to hospital (i.e. I went home without spending a night in hospital) (2)
- Yes, and I was admitted to hospital (i.e. I spent one or more nights in hospital) (3)

| Page Break |
| --- |

Display This Question:

If Did you go to hospital for treatment of your asthma/COPD attack? = Yes, I attended a hospital accident and emergency department but I was not admitted to hospital (i.e. I went home without spending a night in hospital)

Or Did you go to hospital for treatment of your asthma/COPD attack? = Yes, and I was admitted to hospital (i.e. I spent one or more nights in hospital)

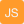

M3.24.1_ud_v3 On what date did you attend hospital?  Please select a date on the calendar below. 

 If you are not sure of the exact date, click on the approximate date.

________________________________________________________________

M4 Since you last checked in with us, have you had any other **serious** illness or injury? 
 
*i.e. something that did NOT present with loss of smell or taste, fever, persistent cough, fatigue, diarrhoea, abdominal pain or loss of appetite or an attack (exacerbation) of asthma or COPD symptoms.*
 
By serious, we mean an illness or injury:
 - that was life-threatening, or
 - that caused you to be admitted to hospital, or
 - that prolonged your hospital stay (if you were already in hospital when it happened), or
 - that resulted in a lasting disability or permanent damage to your health

 We ONLY need to know about **serious** medical events you have experienced since you last checked in with us, i.e. just those which fulfill the criteria above. If you had an illness or injury that does not fullfill the criteria above, click ‘No’ below to move to the next question.

- Yes, I have one or more other serious illnesses or injuries since my last questionnaire (1)
- No, I have not had any other serious illnesses or injuries since my last questionnaire (2)

| Page Break |
| --- |

Display This Question:

If Since you last checked in with us, have you had any other serious illness or injury?    i.e. some... = Yes, I have one or more other serious illnesses or injuries since my last questionnaire

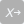

M4.05 How many other **serious** illnesses or injuries have you had since you last checked in with us?

▼ 1 (1) ... 10 (10)

| Page Break |
| --- |

End of Block: COVIDENCE_Recent Health

Start of Block: COVIDENCE_Other illness_ Part 2 and Pregnancy_Part 1

Display This Question:

If Since you last checked in with us, have you had any other serious illness or injury?    i.e. some... = Yes, I have one or more other serious illnesses or injuries since my last questionnaire

M4.1 Please name and briefly describe the nature of any other **serious** illnesses / injuries that you have experienced since your last questionnaire here (e.g. heart attack, stroke).
 
If you have more than one illness or injury to report, put each one in a separate box.

Display This Choice:

If Illnesses >= 1

Display This Choice:

If Illnesses >= 2

Display This Choice:

If Illnesses >= 3

Display This Choice:

If Illnesses >= 4

Display This Choice:

If Illnesses >= 5

Display This Choice:

If Illnesses >= 6

Display This Choice:

If Illnesses >= 7

Display This Choice:

If Illnesses >= 8

Display This Choice:

If Illnesses >= 9

Display This Choice:

If Illnesses >= 10

|  | Illness (4) | Description of illness (5) |
| --- | --- | --- |
| Display This Choice:  If Illnesses >= 1  Illness 1 (1) |  |  |
| Display This Choice:  If Illnesses >= 2  Illness 2 (4) |  |  |
| Display This Choice:  If Illnesses >= 3  Illness 3 (5) |  |  |
| Display This Choice:  If Illnesses >= 4  Illness 4 (6) |  |  |
| Display This Choice:  If Illnesses >= 5  Illness 5 (7) |  |  |
| Display This Choice:  If Illnesses >= 6  Illness 6 (8) |  |  |
| Display This Choice:  If Illnesses >= 7  Illness 7 (9) |  |  |
| Display This Choice:  If Illnesses >= 8  Illness 8 (10) |  |  |
| Display This Choice:  If Illnesses >= 9  Illness 9 (11) |  |  |
| Display This Choice:  If Illnesses >= 10  Illness 10 (12) |  |  |

| Page Break |
| --- |

Display This Question:

If Since you last checked in with us, have you had any other serious illness or injury?    i.e. some... = Yes, I have one or more other serious illnesses or injuries since my last questionnaire

M4.1.1 Please describe the nature of the other **serious** illnesses/injuries that you have experienced since your last questionnaire here.

|  | Was this illness/injury life threatening? | | Were you admitted to hospital for this illness/injury? | |
| --- | --- | --- | --- | --- |
|  | Yes (1) | No (2) | Yes (1) | No (2) |
| ${M4.1/ChoiceTextEntryValue/1/4} (1) |  |  |  |  |
| ${M4.1/ChoiceTextEntryValue/4/4} (4) |  |  |  |  |
| ${M4.1/ChoiceTextEntryValue/5/4} (5) |  |  |  |  |
| ${M4.1/ChoiceTextEntryValue/6/4} (6) |  |  |  |  |
| ${M4.1/ChoiceTextEntryValue/7/4} (7) |  |  |  |  |
| ${M4.1/ChoiceTextEntryValue/8/4} (8) |  |  |  |  |
| ${M4.1/ChoiceTextEntryValue/9/4} (9) |  |  |  |  |
| ${M4.1/ChoiceTextEntryValue/10/4} (10) |  |  |  |  |
| ${M4.1/ChoiceTextEntryValue/11/4} (11) |  |  |  |  |
| ${M4.1/ChoiceTextEntryValue/12/4} (12) |  |  |  |  |

| Page Break |
| --- |

Display This Question:

If Since you last checked in with us, have you had any other serious illness or injury?    i.e. some... = Yes, I have one or more other serious illnesses or injuries since my last questionnaire

M4.1.2 Please describe the nature of the other **serious** illnesses/injuries that you have experienced since your last questionnaire here.

|  | If you were already in hospital when you experienced this illness or injury, did it prolong your hospital stay? | | | Did this illness or injury result in a lasting disability or permanent damage to your health? | |
| --- | --- | --- | --- | --- | --- |
|  | Yes (1) | No (2) | N/A (3) | Yes (1) | No (2) |
| ${M4.1/ChoiceTextEntryValue/1/4} (1) |  |  |  |  |  |
| ${M4.1/ChoiceTextEntryValue/4/4} (4) |  |  |  |  |  |
| ${M4.1/ChoiceTextEntryValue/5/4} (5) |  |  |  |  |  |
| ${M4.1/ChoiceTextEntryValue/6/4} (6) |  |  |  |  |  |
| ${M4.1/ChoiceTextEntryValue/7/4} (7) |  |  |  |  |  |
| ${M4.1/ChoiceTextEntryValue/8/4} (8) |  |  |  |  |  |
| ${M4.1/ChoiceTextEntryValue/9/4} (9) |  |  |  |  |  |
| ${M4.1/ChoiceTextEntryValue/10/4} (10) |  |  |  |  |  |
| ${M4.1/ChoiceTextEntryValue/11/4} (11) |  |  |  |  |  |
| ${M4.1/ChoiceTextEntryValue/12/4} (12) |  |  |  |  |  |

Start of Block: COVIDENCE_Demographics

M9 The next section asks about any changes to your employment and lifestyle since you last completed one of our questionnaires.

| Page Break |
| --- |

M9.1 Has your employment status changed since you last checked in with us?

- Yes (1)
- No (2)

| Page Break |
| --- |

Display This Question:

If Has your employment status changed since you last checked in with us? = Yes

M9.2 Which of the following best describes your current occupational status?

- Employed (1)
- Self-employed (2)
- Retired (3)
- Furloughed (9)
- Unemployed (4)
- Not working due to sickness/ disability or illness (5)
- Never employed (6)
- Student (7)
- Other	(8)

| Page Break |
| --- |

Display This Question:

If Which of the following best describes your current occupational status? = Unemployed

M9.3 How long have you been unemployed?

- Less than 1 month (4)
- 1-3 months (5)
- 3-6 months (6)
- 6-12 months (7)
- 1-4 years (8)
- 5 years or more (9)

| Page Break |
| --- |

M9.10 **Since you last checked in with us,** has your household income been sufficient to cover the basic needs of your household, such as food and heating?

- Yes (17)
- Mostly (18)
- Sometimes (19)
- No (20)

| Page Break |
| --- |

M9.11 **Since you last checked in with us,** have you claimed Universal Credit?

- Yes, I have applied to receive Universal Credit but have **not yet** received any payments (1)
- Yes, I have claimed Universal Credit and received **one or more** payments (2)
- No (3)

End of Block: COVIDENCE_Demographics

Start of Block: COVIDENCE_Lifestyle

M10.1
Which of the following best describes your use of cigarettes over the last month?

- I did not smoke any cigarettes (19)
- I smoked cigarettes occasionally but not everyday (20)
- I smoked cigarettes daily (21)

| Page Break |
| --- |

M10.2 Which of the following best describes your use of e-cigarettes (vaping) over the last month?

- I did not vape or use e-cigarettes (49)
- I smoked vape or used e-cigarettes occasionally but not everyday (50)
- I smoked vape or used e-cigarettes daily (51)

| Page Break |
| --- |

M10.5 During the last week, roughly how many hours did you spend doing light exercise that does not make you particularly breathless, such as light gardening,  walking, including walking for pleasure or exercise, walking to the shops, walking to work?

▼ 0 (28) ... 10+ (38)

| Page Break |
| --- |

M10.6 During the last week, roughly how many hours did you spend doing more vigorous physical exercise of sufficient intensity to make you breathless or to raise your heart rate significantly, such as heavy physical work, more strenuous gardening (e.g. vigorous digging, landscaping) swimming, jogging, aerobics, football, tennis, cycling, gym workout?

▼ 0 (28) ... 10+ (38)

| Page Break |
| --- |

M10.7 During the last week, roughly how many hours did you spend doing lower impact physical exercise to improve flexibility or core strength such as yoga, tai chi or pilates?

▼ 0 (28) ... 10+ (38)

| Page Break |
| --- |

M10.8 How many units of alcohol did you drink over the last 7 days? 

(One unit is a ½ a pint (285 ml) of ordinary beer, lager or cider; 25 ml of spirits; 1 small glass (75 ml) of wine; or 50 ml of sherry).

- None (28)
- 1-7 units (29)
- 8-14 units (30)
- 15-21 units (31)
- 22-28 units (32)
- More than 28 units (33)

| Page Break |
| --- |

M10.11 During the past month, how many hours of actual sleep did you get per night on average? (This may be different than the number of hours you spent in bed)

▼ 0 (1) ... 24 (25)

End of Block: COVIDENCE_Lifestyle

Start of Block: COVIDENCE_Nutritional Supplements

M12 Over the **last month**, have you taken any of the following supplements at least **once per week**? 
Select all that apply.

- Multivitamin (including prenatal multivitamins) (1)
- Supplement containing vitamin A only (2)
- Supplement containing vitamin B only (3)
- Supplement containing vitamin C only (4)
- Supplement containing vitamin D only (5)
- Supplement containing calcium only (6)
- Supplement containing calcium and vitamin D combined (7)
- Supplement containing vitamin E only (8)
- Supplement containing zinc only (9)
- Supplement containing selenium only (21)
- Supplement containing iron only (10)
- Supplement containing probiotics (11)
- Supplement containing fish oil, krill oil or other source of omega-3 fatty acids (12)
- Supplement containing cod liver oil (13)
- Supplement containing echinacea (14)
- Supplement containing garlic or garlic powder (allicin) (15)
- Supplement containing turmeric / curcumin (16)
- Supplement containing Cannabidiol (CBD) oil (18)
- Supplements containing folic acid (19)
- Other (e.g. other micronutrients (such as herbal supplements) or combinations of micronutrients (such vitamin C & zinc)) Please specify: (20) ________________________________________________
- ⊗None of the above (17)

| Page Break |
| --- |

Start of Block: COVIDENCE_General Health

M13 Over the last month, would you say that on the whole, your health has been:

- Excellent	(13)
- Very good (14)
- Good (15)
- Fair (16)
- Poor (17)

| Page Break |
| --- |

Start of Block: COVIDENCE_Quality of Life

M14.1 Which of these statements best describes your breathing currently?

- I get breathless only with strenuous exercise (1)
- I get short of breath when hurrying on level ground or up a slight hill (2)
- I am slower than most people of the same age on a level surface or have to stop when walking at my own pace on the level (3)
- I stop for breath walking 100 metres or after a walking few minutes at my own pace on the level (4)
- I am too breathless to leave the house (5)

| Page Break |
| --- |

M15.3_ud Would YOU say that you currently have 'long COVID', i.e. ongoing symptoms more than four weeks after the onset of proven or suspected COVID-19?

- Yes (1)
- No (2)
- Don't know / not sure (3)

| Page Break |
| --- |

Display This Question:

If Would YOU say that you currently have 'long COVID', i.e. ongoing symptoms more than four weeks af... = Yes

M15.1_ud We will now ask two detailed questions about long-term symptoms which are commonly reported by people who have had COVID. 

 First, have you or your family noticed that you are currently experiencing any of the following problems?

|  | No (1) | Yes but improving (2) | Yes, but not improving or worsening (3) | Yes and worsening (4) |
| --- | --- | --- | --- | --- |
| Excessive shortness of breath (1) |  |  |  |  |
| Coughing (2) |  |  |  |  |
| Unusual tiredness or fatigue (3) |  |  |  |  |
| Problems with sleeping (4) |  |  |  |  |
| Memory problems (5) |  |  |  |  |
| Difficulty concentrating (6) |  |  |  |  |
| Pains in your muscles or joints (7) |  |  |  |  |
| Problems with your sense of smell / taste (8) |  |  |  |  |
| Diarrhoea (9) |  |  |  |  |
| Stomach (abdominal) pains (10) |  |  |  |  |
| Changes to your voice (11) |  |  |  |  |
| Hair loss (12) |  |  |  |  |
| Unusual racing of the heart (13) |  |  |  |  |
| Lightheadedness or dizziness (14) |  |  |  |  |
| Unusual sweating (15) |  |  |  |  |
| Ringing or buzzing in your head or in one or both ears that lasts for more than five minutes at a time (16) |  |  |  |  |

| Page Break |
| --- |

Display This Question:

If Would YOU say that you currently have 'long COVID', i.e. ongoing symptoms more than four weeks af... = Yes

M15.2_ud
Second, here is a list of statements relating to common long-term symptoms of COVID-19. Please select one answer per line to indicate how you have been feeling over the past 7 days.

|  | Not at all (1) | A little bit (2) | Somewhat (3) | Quite a bit (4) | Very much (5) |
| --- | --- | --- | --- | --- | --- |
| I feel fatigued (1) |  |  |  |  |  |
| I feel weak all over (2) |  |  |  |  |  |
| I feel listless ("washed out") (3) |  |  |  |  |  |
| I feel tired (4) |  |  |  |  |  |
| I have trouble starting things because I am tired (5) |  |  |  |  |  |
| I have trouble finishing things because I am tired (6) |  |  |  |  |  |
| I have energy (7) |  |  |  |  |  |
| I am able to do my usual activities (8) |  |  |  |  |  |
| I need to sleep during the day (9) |  |  |  |  |  |
| I am too tired to eat (10) |  |  |  |  |  |
| I need help doing my usual activities (11) |  |  |  |  |  |
| I am frustrated by being too tired to do the things I want to do (12) |  |  |  |  |  |
| I have to limit my social activity because I am tired (13) |  |  |  |  |  |

| Page Break |
| --- |

HEAD By selecting one box in each group below, please indicate which statements best describe your own health state today.

M14.2 Mobility

- I have no problems in walking about (1)
- I have some problems in walking about (2)
- I am confined to bed (3)

| Page Break |
| --- |

M14.3 Self-Care

- I have no problems with self-care (1)
- I have some problems washing or dressing myself (2)
- I am unable to wash or dress myself (4)

| Page Break |
| --- |

M14.4 Usual Activities (e.g. work, study, housework, family or leisure activities)

- I have no problems with performing my usual activities (1)
- I have some problems with performing my usual activities (5)
- I am unable to perform my usual activities (4)

| Page Break |
| --- |

M14.5 Pain / Discomfort

- I have no pain or discomfort (1)
- I have moderate pain or discomfort (2)
- I have extreme pain or discomfort (3)

| Page Break |
| --- |

M14.6 Anxiety / Depression

- I am not anxious or depressed (1)
- I am moderately anxious or depressed (2)
- I am extremely anxious or depressed (3)

End of Block: COVIDENCE_Quality of Life

Start of Block: June_COVIDENCE_Your own health state today

Q364_horz We would like to know how good or bad your health is TODAY.    You will see a scale numbered from 0 to 100.  100 means the best health you can imagine.  0 means the worst health you can imagine.    Please move the slider to indicate on the scale how your health state is TODAY.

|  | **The worst health you can imagine** | **The best health you can imagine** |
| --- | --- | --- |

|  | 0 | 10 | 20 | 30 | 40 | 50 | 60 | 70 | 80 | 90 | 100 |
| --- | --- | --- | --- | --- | --- | --- | --- | --- | --- | --- | --- |

| () | 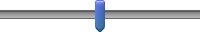 |
| --- | --- |

| Page Break |
| --- |

End of Block: June_COVIDENCE_Your own health state today

Start of Block: COVIDENCE_Mental well-being

M16.1 We would now like to ask a few questions about your mental well-being.
 
Our mental health, particularly depression and anxiety, is closely entwined with our physical health and can play a role in how well we fight an illness or respond to treatment. That’s why we want to ask these questions.

 In the last week, how often have you been bothered by the following problems

|  | Not at all (1) | Several days (1-3 days/week) (2) | More than half the days (4-5 days/week) (3) | Nearly every day (6-7 days/week) (4) |
| --- | --- | --- | --- | --- |
| Little interest or pleasure in doing things (1) |  |  |  |  |
| Feeling down, depressed, or hopeless (2) |  |  |  |  |
| Feeling nervous, anxious or on edge (3) |  |  |  |  |
| Not being able to stop or control worrying (5) |  |  |  |  |

| Page Break |
| --- |

End of Block: COVIDENCE_Mental well-being

M18.2 In the last week, how many journeys did you make on public transport? (return trips count as 2 journeys)

▼ 0 (1) ... 10 or more (15)

| Page Break |
| --- |

M18.3 In the last week, how frequently have you worn a face mask or face covering while in an indoor public place that is outside of work, e.g. in shops, on public transport etc?

- Always (100% of the time) (1)
- Usually (50%-99%) (2)
- Sometimes (1-49% of the time) (3)
- Never (0% of the time) (4)
- Not applicable (I haven't been in a public place in the last week) (5)

| Page Break |
| --- |

Display This Question:

If In the last week, how frequently have you worn a face mask or face covering while in an indoor pu... = Always (100% of the time)

Or In the last week, how frequently have you worn a face mask or face covering while in an indoor pu... = Usually (50%-99%)

Or In the last week, how frequently have you worn a face mask or face covering while in an indoor pu... = Sometimes (1-49% of the time)

M18.31 What type of face mask or face covering did you most frequently use in the last week while in an indoor public place outside of work, e.g. in shops, on public transport etc? 

Please select one option - if you used more than one type of mask in the last week, select the one that you used most frequently. 

 (For information, the abbreviations FFP2 and N95 below refer to masks that are designed to offer enhanced protection: they will usually bear a label (e.g. CE mark or UKCA mark) to indicate that they have been certified as providing a given level of protection).

- Disposable surgical mask (not FFP2- or N95-certified) (1)
- Simple cloth face covering (not FFP2- or N95-certified) (2)
- Disposable FFP2 or N95 mask (3)
- Reusable FFP2 or N95 cloth face mask (4)
- Other: please specify (5) ________________________________________________

| Page Break |
| --- |

M18.6 Over the last week, how many times have you been visited at home by someone who does not live in your household? 

(We are referring to visitors entering inside your home. People who just call at the door and do not come inside do not count)

▼ 0 (16) ... 10 or more (15)

M18.7 Over the last week, how many times have you visited someone who does not live in your house? 

(We are referring to visits in which you enter inside someone else’s home. Visits where you do not cross the threshold do not count. Neither do visits to the shops or other public places, we will ask about these later.)

▼ 0 (16) ... 10 or more (15)

| Page Break |
| --- |

M18.8 In the last week, how many times have you been inside a shop or supermarket?

▼ 0 (16) ... 10 or more (15)

| Page Break |
| --- |

M18.9 In the last week, how often have you been inside another indoor public space (e.g. café, pub, place of worship, restaurant, gym, day centre, waiting room, school, library, entertainment venue, hairdresser, takeaway restaurant)?

▼ 0 (16) ... 10 or more (15)

| Page Break |
| --- |

M18.92 Since you last checked in with us, have you travelled outside of the UK?

- Yes (1)
- No (2)

| Page Break |
| --- |

Display This Question:

If Since you last checked in with us, have you travelled outside of the UK? = Yes

M18.92.1 If yes, please write country or countries visited here

________________________________________________________________

________________________________________________________________

________________________________________________________________

________________________________________________________________

________________________________________________________________

| Page Break |
| --- |

M20 Is there any other information you think would be useful to us, or any clarification you would like to make regarding the information you have provided today?

- Yes (1)
- No (2)

| Page Break |
| --- |

Display This Question:

If Is there any other information you think would be useful to us, or any clarification you would li... = Yes

M20.1 If yes, please provide details:

________________________________________________________________

________________________________________________________________

________________________________________________________________

________________________________________________________________

________________________________________________________________

End of Block: COVIDENCE_Response to Coronavirus Outbreak_continued
